# Vitamins D_2_ and D_3_ have overlapping but different effects on human gene expression revealed through analysis of blood transcriptomes in a randomised double-blind placebo-controlled food-fortification trial

**DOI:** 10.1101/2020.12.16.20247700

**Authors:** Louise R. Durrant, Giselda Bucca, Andrew Hesketh, Carla Möller-Levet, Laura Tripkovic, Huihai Wu, Kathryn H. Hart, John C. Mathers, Ruan M. Elliott, Susan A. Lanham-New, Colin P. Smith

## Abstract

For the first time, we report the influence of vitamin D_2_ and vitamin D_3_ on genome-wide gene expression in whole blood from healthy women representing two ethnic groups, white European and South Asian. In this randomised placebo-controlled trial, participants were given daily physiological doses (15 µg) of either vitamin D_2_ or D_3_ for 12 weeks and changes in the transcriptome were compared relative to the transcriptome at baseline. While there was some overlap in the repertoire of differentially expressed genes after supplementation with each vitamin D source, most changes were specific to either vitamin D_3_ or vitamin D_2_, suggesting that each form of the vitamin may have different effects on human physiology. Notably, following vitamin D_3_ supplementation, the majority of changes in gene expression reflected a down-regulation in the activity of genes, many encoding components of the innate and adaptive immune systems. These are consistent with the emerging concept that vitamin D orchestrates a shift in the immune system towards a more tolerogenic status. Moreover, divergent changes were observed following supplementation with either vitamin D_3_ or vitamin D_2_ for gene expression associated with type 1 and type 2 interferon activity. This is particularly intriguing as interferons play a critical role in the innate response to infection and aberrant type 1 interferon signalling is implicated in severe COVID-19 disease. The observed differences in gene expression after supplementation with vitamin D_2_ compared with vitamin D_3_ warrant a more intensive investigation of the biological effects of the two forms of vitamin D on human physiology.

**Significance statements:** This study suggests that the influence of vitamins D_2_ and D_3_ on human physiology may not be the same, as deduced from differences in gene expression within whole blood.

South Asian participants were found to respond differently to vitamin D supplementation at the transcriptome level from white Europeans.

The differentially expressed immune pathways identified in this study are consistent with vitamin D orchestrating a more tolerogenic immune status and this could be relevant in the context of the severity of immune response to viral infections such as Covid-19.

The potential relevance of this study to severe Covid-19 disease is highlighted by our observed enhancement of type 1 interferon signalling by vitamin D_3_, but not vitamin D_2_.

## Introduction

Vitamin D is a pro-hormone that is critical for good health. While vitamin D’s function in maintaining the musculo-skeletal system is best understood, it fulfils myriad roles in human physiology, including maintaining a healthy immune system (Prietl et al., 2013, Martens et al., 2020). Vitamin D deficiency (defined as serum concentrations of 25-hydroxyvitamin D (25(OH)D) below 25 nmol/L) or inadequacy (defined as serum level of 25(OH)D below 50 nmol/L) is considered a global pandemic and a public health issue of great importance in the human population, especially in older people, individuals not exposed to sufficient sunlight and, importantly, ethnic minority groups with dark skin tone (Kohlmeier, 2020, Lanham-New et al., 2020). Vitamin D deficiency is considered to have many detrimental effects on human health which include increased risk of osteoporotic and stress fractures, increased risk of developing cardiovascular diseases and some cancers, and poor modulation of the immune system (Holick, 2007, Bouillon, 2018). Low vitamin D status is associated with higher mortality, including death from cancer (Zhang et al., 2019).

Vitamin D_3_ (cholecalciferol) is produced in the skin by the action of ultraviolet B radiation from the sun. Following two successive hydroxylation steps, in the liver and kidney, respectively, the active form of the vitamin, 1,25-dihydroxyvitamin D_3_ (1,25(OH)D_3_) binds to the intracellular vitamin D receptor (VDR) and, in complex with the retinoid X receptor (RXR), the heterodimer regulates expression of hundreds of genes through the vitamin D response element (VDRE) (Long et al., 2015). Vitamin D can also be obtained from foods or as a supplement. Both the plant/fungus-derived vitamin D_2_ (ergocalciferol) and the animal-derived D_3_ forms are available as nutritional supplements. Both can be hydroxylated into their active forms and bind to the VDR with similar affinity, but there are differences in their catabolism and in their binding affinity to vitamin D binding protein (DBP), the major vitamin D transport protein in blood. Vitamin D_2_ binds DBP with lower affinity and is catabolised faster (Hollis, 1984, Houghton and Vieth, 2006).

The functional equivalence, or otherwise, of vitamins D_2_ and D_3_ for human health has been a subject of much debate in recent years, with some authors suggesting that the two compounds have equal efficacy while others provide evidence that vitamin D_3_ increases circulating serum 25(OH)D concentration more efficiently than vitamin D_2_ (Tripkovic et al., 2012, Wilson et al., 2017, Biancuzzo et al., 2010a, Biancuzzo et al., 2010b, Lanham-New et al., 2010). Current guidance given to the general public by the US National Institutes of Health (NIH), the UK Department of Health and Social Care, and other government bodies around the world lack acknowledgement that the two forms of vitamin D are not considered equivalent or equally effective when used at nutritional (physiological) doses, while at high doses vitamin D_2_ is less potent. The NIH state that most evidence indicates that vitamin D_3_ increases serum 25(OH)D levels to a greater extent and maintains these higher levels longer than vitamin D_2_, even though both forms are well absorbed in the gut (ODS, 2020).

Although vitamin D is best known for its role in maintaining bone health and calcium homeostasis, it also exerts a broad range of extra-skeletal effects on cellular physiology and on the immune system (Pilz et al., 2019, Schwalfenberg, 2011, Christakos et al., 2016, Rosen et al., 2012) and multiple studies have linked poor vitamin D status with the pathogenesis of immune mediated inflammatory diseases (Damoiseaux and Smolders, 2018). Vitamin D is essential for human health and it is recommended that individuals maintain serum concentrations of at least 25 nmol/L of 25(OH)D (the accepted biomarker of systemic vitamin D status), throughout the year and throughout the life course (SACN, 2016). However, it can be difficult to sustain such levels of serum 25(OH)D year-round from diet and sunlight alone, particularly amongst ‘at-risk’ groups, such as those with limited sun exposure or those with darker skin tone living in northern latitudes (above 40°N).

In acute studies comparing the efficacy of vitamins D_2_ and D_3_ in raising serum 25(OH)D concentration, vitamin D_2_ is less effective than D_3_ when given as single bolus (Tripkovic et al., 2012). However, the findings from studies following the daily administration of D_2_ or D_3_ over longer time-periods are more equivocal, with clinical trials showing higher efficacy of D_3_ (Zittermann et al., 2014, Tripkovic et al., 2017) or equal efficacy (Holick et al., 2008). In a recent meta-analysis, supplementation with vitamins D_2_ and D_3_ were found equally effective in raising 25(OH)D concentrations in infants up to 1 year of age (Zittermann et al., 2020). It should be noted that supplementation with one form of vitamin D can reduce circulating concentrations of the other form. For example, our recent comparative trial of D_2_ versus D_3_ revealed that serum 25(OH)D_3_ concentration was reduced over the 12-week intervention in participants given vitamin D_2_, compared with the average 12-week concentration in participants given placebo ((Tripkovic et al., 2017), see Fig. 1C, for example). The implications of such reciprocal depletion remain to be explored.

**Figure 1.**
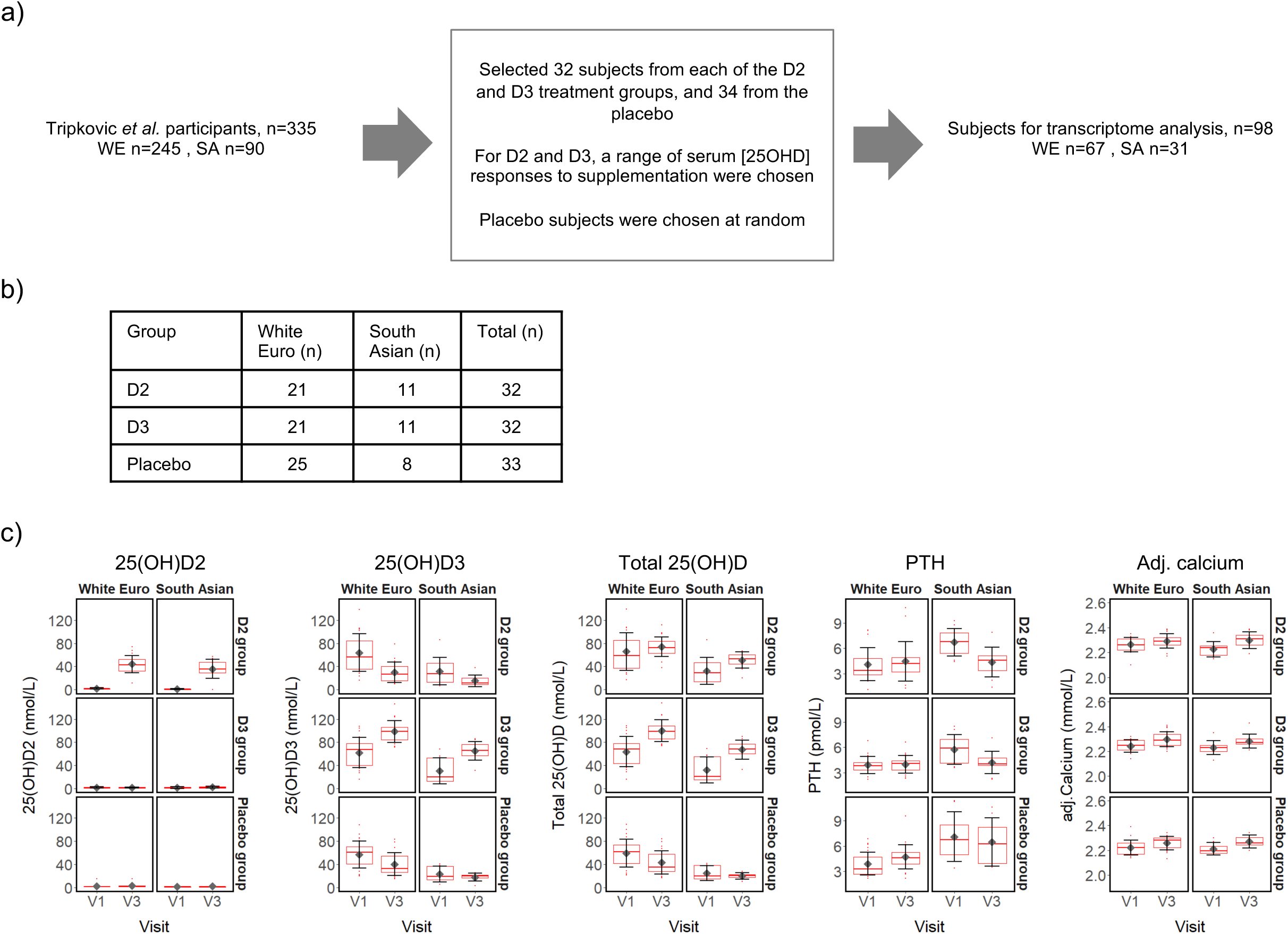
(a) Selection of 98 subject participants from the Tripkovic et al. (2017) study (Tripkovic et al., 2017) for transcriptomic analysis of their V1 (baseline) and V3 (12-week) samples in the present study; (b) Ethnicity and treatment group membership for the 97 subjects for which transcriptomic data was obtained (data for one South Asian subject from the placebo group did not pass quality control); (c) Metadata on serum concentrations of 25(OH)D_2_, 25(OH)D_3_, total 25(OH)D, PTH and calcium (albumin-adjusted) for the 97 subjects (see **Supplementary Data File 1** for details). Participants were selected to provide comparable numbers between the placebo and the two vitamin D treatment groups, covering the full range of serum responses to supplementation within the D_2_ and D_3_ treatment groups, as judged from the measured changes in serum 25(OH)D_2_ or 25(OH)D_3_ concentrations between V1 and V3.

There is limited understanding of the effects of vitamin D supplementation on gene expression *in vivo* in humans because of the diversity of experimental designs used. This includes: (i) diverse sampling intervals, ranging from hours to years following vitamin D supplementation (Berlanga-Taylor et al., 2018, Neme et al., 2019); (ii) use of substantially different doses ranging from physiological (moderate) to supra-physiological doses; and (iii) relatively small numbers of participants were recruited so that most studies were considerably underpowered (Neme et al., 2019, Hossein-nezhad et al., 2013). Currently, there is no robust evidence, from *in vivo* human genome-wide expression analysis, about which specific cellular pathways are influenced by vitamin D supplementation. Moreover, the influence of vitamin D_2_, as distinct from vitamin D_3_, on gene expression in humans has not yet been evaluated, even though vitamin D_2_ is used widely as a supplement and food fortificant.

We have addressed this gap in knowledge by investigating gene expression in a relatively large cohort of healthy white European and South Asian women who participated in a randomised double-blind placebo-controlled trial that compared the relative efficacy of vitamins D_2_ and D_3_ in raising serum 25(OH)D concentration. The study concluded that vitamin D_3_ was superior to vitamin D_2_ in raising serum 25(OH)D concentration (Tripkovic et al., 2017). As an integral part of the original study design, we also investigated gene expression in the participants over the 12-week trial period. This allowed us to examine the effects of vitamin D supplementation on the transcriptome and to determine whether these effects might differ depending on supplementation with vitamin D_2_ compared with vitamin D_3_. We therefore sampled blood at the beginning, middle and end of the 12-week intervention and quantified changes in the whole blood transcriptome. Surprisingly, we observed that the two vitamins exerted overlapping but different effects on the human blood transcriptome. Our findings support the hypothesis that the biological effects of vitamin D_2_ and D_3_ may differ in humans and suggest that a more comprehensive analysis of the biological effects of the two forms of vitamin D on human physiology is warranted. In this context, it is important to note that a recent meta-analysis of vitamin D supplementation trials found that reduced cancer mortality was seen only with vitamin D_3_ supplementation, not with vitamin D_2_ supplementation, and indicated that all-cause mortality was significantly lower in trials with vitamin D_3_ supplementation than in trials with vitamin D_2_ (Zhang et al., 2019).

## Results

The study volunteers were South Asian (SA) and white European (WE) women based in the United Kingdom, aged between 20 and 64 years (n=335; (Tripkovic et al., 2017)) and participation was for 12 weeks over the winter months (October to March, in Surrey, UK; latitude, 51°14’ N). Serum measurements, including concentrations of total 25(OH)D, 25(OH)D_2_ and 25(OH)D_3_, were determined from fasting blood samples taken at the start (baseline defined as Visit 1 (V1)) and at weeks 6 (V2) and 12 (V3). To determine whether changes in serum 25(OH)D concentration as a result of vitamin D supplementation are associated with physiological changes at the level of gene expression, we investigated the whole blood transcriptome from a representative subset of the study participants (n=98) using total RNA isolated from V1 and V3 blood samples and Agilent Human Whole Genome 8× 60K v2 DNA microarrays (Fig. 1). Participants for transcriptome analysis were selected to provide comparable numbers between the placebo and the two vitamin D treatment groups, covering the full range of serum responses to supplementation within the D_2_ and D_3_ treatment groups, as judged from the measured changes in serum 25(OH)D_2_ or 25(OH)D_3_ concentration between V1 and V3 (**Supplementary Data File 1**).

### Metadata for the transcriptome analysis cohort

Following microarray data quality control, transcriptome data were available for both the V1 and V3 samples of 97 study participants (Fig. 1b); the RNA from one participant failed quality control and was excluded. Similar to data reported previously for the entire study cohort (Tripkovic et al., 2017), the sub-set of participants in the present study showed increased concentrations of 25(OH)D_2_ and 25(OH)D_3_ only after supplementation with vitamin D_2_ and vitamin D_3_, respectively, and higher total 25(OH)D in both treatment groups (Fig. 1c). In the vitamin D_3_ treatment groups the mean 25(OH)D_3_ serum levels rose by 59% (WE) and 116% (SA) over the 12-week intervention. Conversely, mean serum 25(OH)D_3_ concentrations fell across the 12 weeks of the study in the placebo groups who did not receive additional vitamin D; 25(OH)D_3_ baseline vitamin D levels in the SA ethnic group tended to be lower than for the WE group and dropped, respectively, by 23% and 29% relative to baseline V1 (Fig. 1c; **Supplementary Data File 1**). It is notable that, in the vitamin D_2_ treatment group, serum 25(OH)D_3_ concentration decreased to a greater extent over the 12-week period, by 52% (SA) and 53% (WE); the implications of this are considered further in the Discussion. Serum 25(OH)D_2_ was low in the absence of specific supplementation, typically less than 5 nmol/L (152/162 samples analysed). Serum calcium concentration, appropriately adjusted for serum albumin concentration, was maintained within a normal clinical range across all sample groups, while the concentration of parathyroid hormone (PTH) was stable between V1 and V3 only within the SA placebo group, the WE vitamin D_2_ and WE vitamin D_3_ intervention groups. PTH concentrations showed an increase at V3 compared with V1 in the WE placebo group, but a decrease in both the vitamin D_2_ and D_3_ intervention groups in the SA ethnic cohort.

### Effects of supplementation with either vitamin D_2_ or D_3_ on global gene expression

Filtering of the normalised microarray data to select for probes showing signals at least 10% above background in at least 41 (∼20%) of the arrays yielded transcript abundance data from 20,662 probes corresponding to 17,588 different genomic features (12,436 of which were annotated with an ENTREZ gene identifier). Using the filtered data, we identified significant differences in the transcriptional responses occurring across the 12-week V1 to V3 period of the study *between* the vitamin D_2_, vitamin D_3_ and placebo treatment groups for each ethnic cohort and, in a separate analysis, to determine significant changes *within* each group between the V3 and V1 sampling points (Fig. 2; **Supplementary Data File 2**). The former was tested in a ‘difference-in-difference’ analysis according to the generalised null hypothesis [treatment1.V3 – treatment1.V1] - [treatment2.V3 – treatment2.V1] = 0 (Fig. 2a, rows 7-12), and the latter using treatment.V3 – treatment.V1 = 0 (Fig. 2a, rows 1-6).

**Figure 2.**
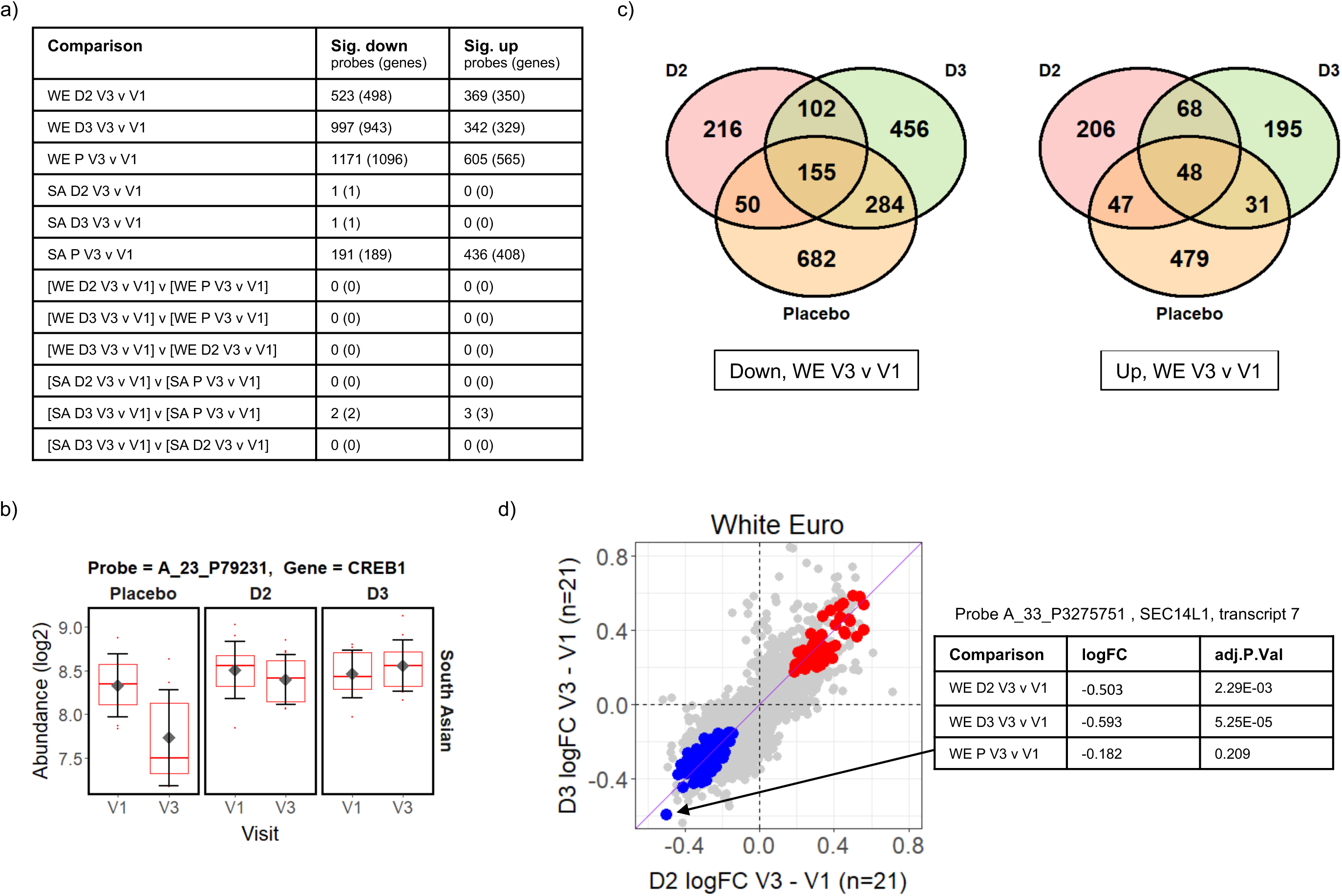
(a) Summary of limma differential expression test results identifying significant changes (adj.P.Val<=0.05) in transcript probe abundance within and between the experimental groups (see **Supplementary Data File 2** for full details). Both the number of unique microarray probes, and the number of unique genome loci they represent, are indicated. WE = “white European”; SA = “South Asian”; D2 = vitamin D_2_ treatment group; D3 = vitamin D_3_ treatment group; P = placebo group. (b) The CREB1 gene probe signal is significantly different (adj.P.Val=0.021) between the vitamin D_3_-treated group and the placebo group over the course of the V1 to V3 study period in the South Asian cohort (i.e. [SA D3 V3 v V1] v [SA P V3 v V1] comparison from Fig. 2a). (c) Venn diagrams showing the numbers of probes in the white European (WE) cohort that are specifically significantly (adj.P.Val<=0.05) down- or up-regulated in V3 compared to V1 in each treatment group. (d) Comparison of the log_2_ fold-changes (FC) in abundance occurring from V1 to V3 in the WE cohort in the D_2_ and D_3_ treatment groups. Probes significantly up-regulated in both D_2_ and D_3_ groups but not the placebo are coloured red, while those similarly down-regulated are shown in blue (see **Supplementary Data File 3**). A probe detecting *SEC14L1* shows the largest decreases in abundance in those down-regulated by D_2_ and D_3_ but not placebo.

No significant changes in probe signals (adj.P.Val<=0.05) were identified in the ‘difference-in-difference’ analysis for the data for the WE ethnic group, but five were found in the corresponding analysis of the SA cohort (Fig. 2a). Furthermore, difference-in-difference analysis of the entire study data, ignoring ethnicity, did not yield statistically significant changes between the D_2_ or D_3_ treatment groups and the placebo control (data not shown). We note that another ‘difference-in-difference’ study on the influence of vitamin D on gene expression (the BEST-D trial) failed to identify any significant changes in gene expression following long-term vitamin D treatment in a Caucasian cohort (Berlanga-Taylor et al., 2018). The observed log_2_ fold-change (log_2_FC) in signal abundances between V3 and V1 in the study typically fall well within +/− 1, and in such a context it is likely that the sample sizes used within the study restrict our ability to reliably identify individual gene expression responses using this highly conservative approach. We consider that inter-individual differences in human populations are likely to hinder detection of small statistically significant gene expression changes *across* treatment groups from microarray-derived data; however, statistically significant differences are detected when evaluating gene expression changes *within* an individual across time (as described below where we examine paired observations with two time points per subject). The five significant changes that were identified as occurring in response to treatment of the SA ethnic group with vitamin D_3_, relative to the placebo, are summarised in **Supplementary Fig. 1**. These are driven primarily by the marked changes in the placebo group between the V1 and V3 sampling points, and may be associated with the sustained low serum 25(OH)D_3_, or high PTH, concentrations observed in this group of subjects. One of these differentially expressed genes encodes the cAMP response element binding protein CREB1 (Fig. 2b) which is part of the cAMP-PKA-CREB signaling pathway in bone cells which contributes to the regulation of skeletal metabolism in response to PTH concentrations (Zhang et al., 2011).

In contrast to the difference-in-difference analysis, the direct *within*-group comparisons of transcript abundance measurements at V3 versus V1 per subject identified large numbers of significant changes in gene expression, most notably in the WE D_2_, WE D_3_ and WE placebo groups (Fig. 2a and c, **Supplementary Data File 2**). These significant gene expression changes in the WE ethnic cohort arising in the vitamin D treatment groups are considered in more detail below. While there is some overlap in the groups of differentially expressed genes, only 13% of down-regulated differentially expressed genes (102 of 774) were common between the two vitamin D treatment groups, while 28% (216 of 774) and 59% (456 of 774) were uniquely down-regulated by D_2_ and D_3_, respectively (excluding those additionally down-regulated in the placebo group over the equivalent 12-week intervention)(Fig 2c). As previously noted above, the serum 25(OH)D_3_ concentration fell over the 12-week intervention period in the vitamin D_2_-treated group; thus, changes in gene expression observed in this group could *a priori* either be due to the influence of vitamin D_2_ itself, or could be attributable to the depletion of the endogenous 25(OH)D_3_ reserves. It was therefore important to have included comparative transcriptomic analysis of the placebo group in this study to distinguish gene expression changes resulting from vitamin D_2_ *per se* from those changes arising due to the depletion of 25(OH)D_3_ levels in this vitamin D_2_ treatment group.

It is notable that expression of a large number of genes was altered between the first and last visits (V1 to V3) in the non-treated placebo group. While some of these changes will be attributable to the effects of the vitamin D depletion observed in this group over the course of the study, there is significant over-representation of genes known to exhibit seasonal differences in gene expression (Dopico et al., 2015) (**Supplementary Fig. 2**). There is however no unique association of seasonally expressed genes with the data for the WE placebo group; a similar significant enrichment is also observed for the significant changes in gene expression identified in the WE D_2_ and WE D_3_ treatment groups (**Supplementary Fig. 2**) and therefore it is unlikely that this apparent seasonal effect exclusively reflects vitamin D-specific effects. There was no overlap between genes significantly down-regulated exclusively in the placebo group and those significantly up-regulated exclusively in the vitamin D treatment groups (or vice versa). Consistent with the 12-week period of the study, seasonal changes in gene expression are therefore a background feature of all the data collected, and is therefore a factor that needs to be considered when undertaking such time course studies.

In the group of probe signals with significantly reduced abundance in the V3 samples compared with V1 in the data for the vitamin D_2_ and D_3_ supplemented participants, but not in the placebo participants, Probe A_33_P3275751, detecting *SEC14L1* gene transcription showed the largest decrease in response to both forms of vitamin D (Fig. 2d). This probe was originally designed to detect ‘transcript variant 7’ and hybridizes to a location at the 3’ end of the *SEC14L1* locus, detecting several splice variants. High expression of *SEC14L1* is significantly associated with lymphovascular invasion in breast cancer patients, where transcript abundance correlates positively with higher grade lymph node metastasis, and poor prognostic outcome (Sonbul et al., 2018). Its overexpression is also frequent in prostate cancer where *SEC14L1* has been identified as a potential biomarker of aggressive progression of the disease (Agell et al., 2012, Burdelski et al., 2015).

### Genes significantly down-regulated by both D_2_ and D_3_, but not placebo, are enriched for functions associated with immune responses

Given the relatively small sample sizes used in this study it is considered more appropriate to examine enrichment of cellular pathways among differentially expressed genes rather than focusing on changes in individual genes. Functional enrichment analysis of the genes represented by the probes specifically repressed by vitamins D_2_ and D_3_ (blue data points in Fig. 2d) suggests that both supplements have suppressive effects on the immune response in the WE ethnic group (Fig. 3 and **Supplementary Data File 3**). Indeed, a significant (p = 1.98E-06) protein-protein interaction network derived from the *Homo sapiens* medium confidence interactions curated in the STRING database (Szklarczyk et al., 2017b) is centred on a highly connected group of proteins associated with the innate immune response, neutrophil degranulation and leukocyte activation (Fig. 3a; all protein groups are detailed in **Supplementary Data File 3**). This includes the histone acetyltransferase, EP300, known to function as a transcriptional coactivator with VDR, the vitamin D receptor protein (Fetahu et al., 2014). Collectively, these findings are consistent with the emerging view that vitamin D exposure leads to a shift from a pro-inflammatory to a more tolerogenic immune status (Prietl et al., 2013, Martens et al., 2020).

**Figure 3.**
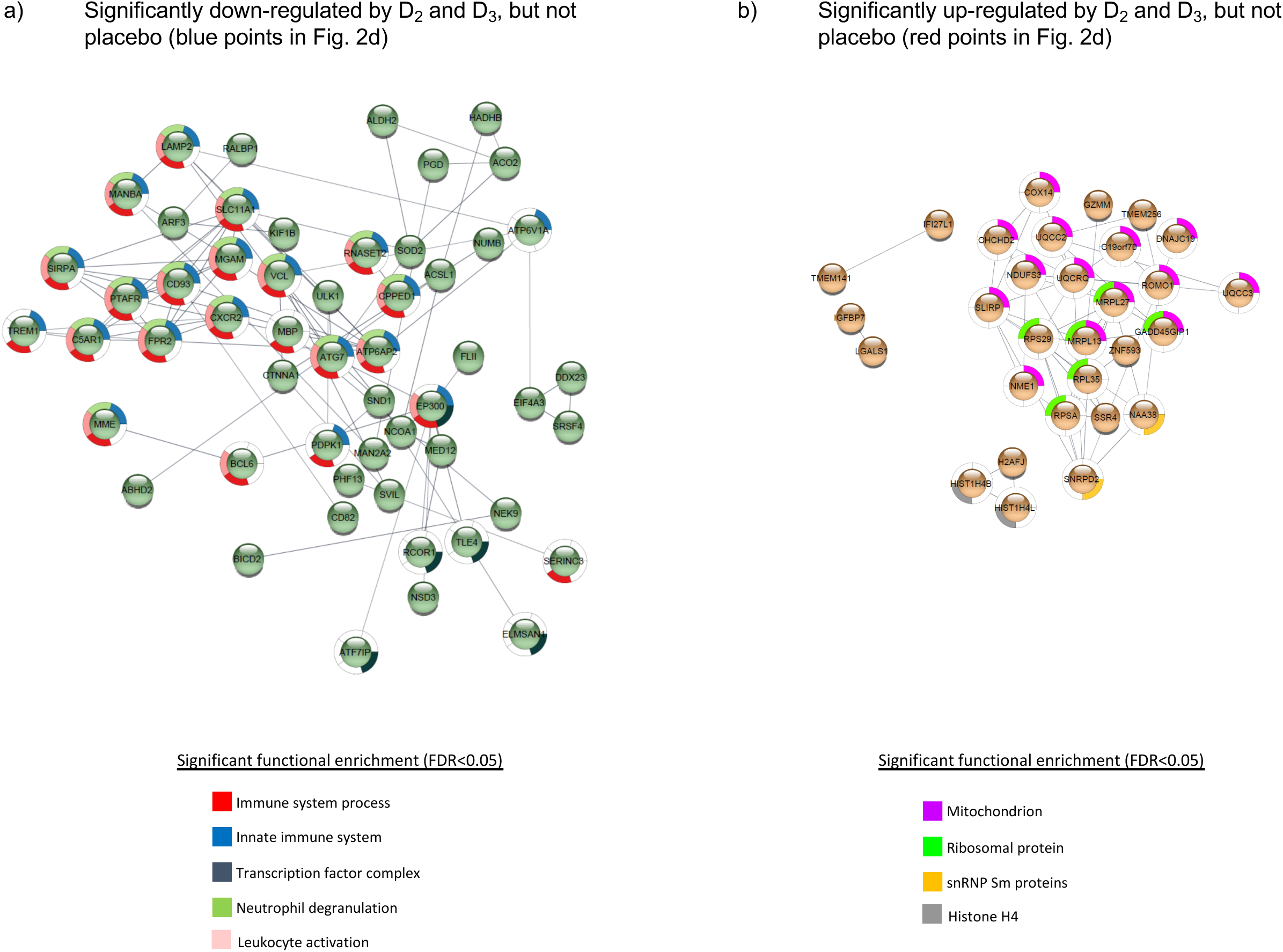
Protein-protein interaction networks for gene products corresponding to the probes (a) significantly down-regulated or (b) significantly up-regulated in both the D_2_ and D_3_ treatment groups of the WE cohort, but not the placebo group. Details given in **Supplementary Data File 3**. The networks were generated using the STRING database of *Homo sapiens* medium confidence (0.4) interactions, and only connected nodes are shown. Networks for both (a) and (b) are significantly enriched for interactions compared to randomised sets, yielding p-values of 1.98 × 10^-6^ and × 10^-9^, respectively.

### Genes significantly up-regulated by both D_2_ and D_3_, but not placebo, include components of histone H4 and the spliceosome

A similar analysis of the probes specifically induced by D_2_ and D_3_ (red points in Fig. 2d) produced a significant (p = 4.99E-09) protein-protein interaction network enriched primarily for mitochondrial and ribosomal proteins, but also involving two subunits of histone H4 and SNRPD2, a core component of the SMN-Sm complex that mediates spliceosomal snRNP assembly (Fig. 3b).

### Comparative functional enrichment analysis supports roles for vitamins D_2_ and D_3_ in the suppression of immunity, and in chromatin modification and spliceosome function

As a complementary approach to the functional enrichment analysis of specific subsets of genes whose expression is altered by supplementation with vitamin D_2_ and D_3_ but not by placebo treatment, a comparative functional enrichment analysis of all significant changes in each treatment group was performed (see Methods). This aimed to identify functional categories that are more extensively affected by vitamin D_2_, or by vitamin D_3_, or by both vitamins D_2_ and D_3_, than by the placebo, and took into consideration the changes occurring in the placebo treatment group across the 12 weeks of the study (Figs. 4 and 5; **Supplementary Data File 4**). Consistent with the earlier analysis, functional categories associated with immunity and immune response pathways are prominent among those genes repressed by vitamin D supplementation (Fig. 4), while mitochondrial, ribosomal and spliceosomal functions are prominent in the induced genes (Fig. 5). Although the two vitamin treatments share many common categories identified from this analysis, the results also highlight some differences between the respective effects of D_2_ or D_3_ supplementation. For example, ‘histone exchange’ is significant only in the vitamin D_2_ up-regulated genes, and ‘chromatin modifying enzymes’ are significant only in the D_3_ down-regulated genes.

**Figure 4.**
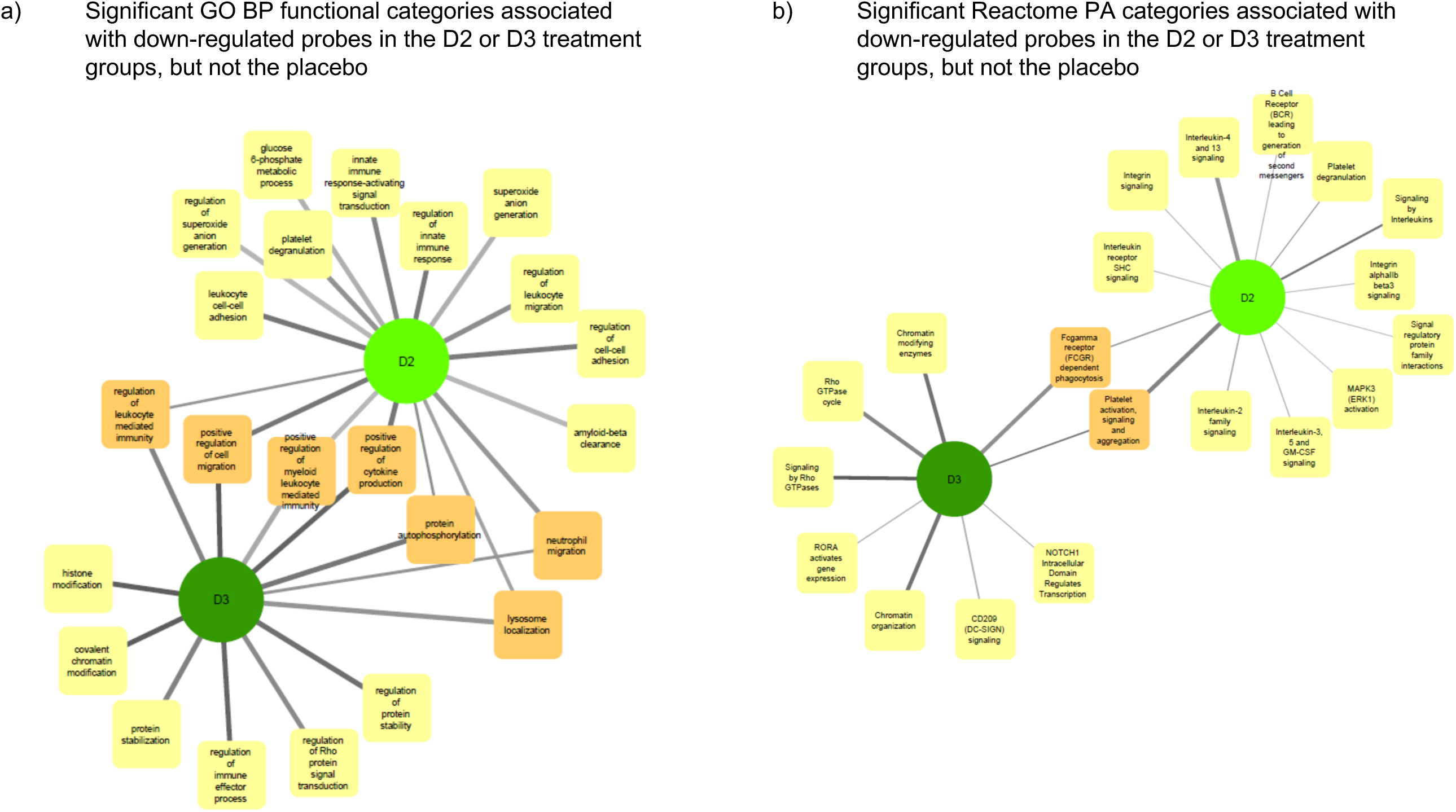
Networks illustrating the functional categories significantly enriched in the gene products represented by the probes significantly down-regulated (adj.P.Val ≤ 0.05) in the D_2_ and D_3_ treatment groups of the WE cohort, but not in the placebo group. Gene products represented by the significantly down-regulated probes in the comparisons WE D2 V3 v V1, WE D3 V3 v V1 and WE P V3 v V1 from Fig. 2(a), and possessing ENTREZ identifiers, were subjected separately to functional enrichment analysis using compareCluster (Yu et al., 2012). The details for each group are given in **Supplementary Data File 4**. Significantly enriched categories (p.adjust ≤ 0.01) from all groups were imported into Cytoscape (Shannon et al., 2003) to visualise categories identified from the D_2_ or D_3_ treatment groups but not by the placebo (see Methods for details). The thickness of lines (edges) drawn to connect the nodes shown increases with increasing significance (p.adjust-values) and only categories where at least one edge is ≤ 1 × 10^-3^ are shown. The complete networks for each differentially expressed group of genes are shown in **Supplementary Data File 5.**

**Figure 5.**
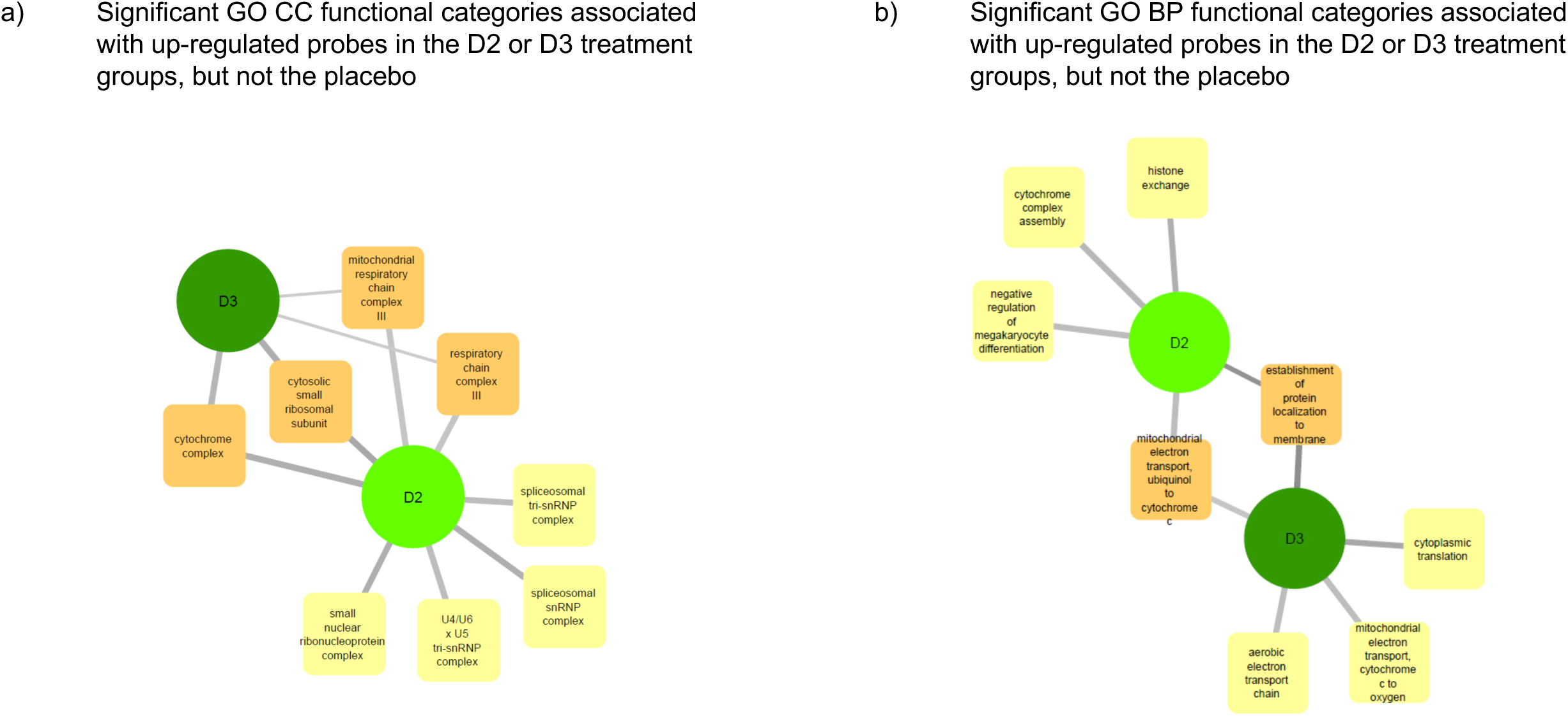
Networks illustrating the functional categories significantly enriched in the gene products represented by the probes significantly up-regulated (adj.P.Val ≤ 0.05) in the D_2_ and D_3_ treatment groups of the WE cohort, but not in the placebo group. Gene products represented by the significantly up-regulated probes in the comparisons WE D2 V3 v V1, WE D3 V3 v V1 and WE P V3 v V1 from Fig. 2(a), and possessing ENTREZ identifiers, were subjected separately to functional enrichment analysis using compareCluster as detailed in the legend to Fig. 4. The complete networks for each differentially expressed group of genes are shown in **Supplementary Data File 5.**

Overall, the observed differences in gene expression from the blood transcriptome presented in this study suggest that the physiological effects of vitamin D_3_ and D_2_ may be dissimilar.

### Weighted gene correlation network analysis (WGCNA) identifies modules of co-expressed genes that significantly correlate with serum markers of vitamin D supplementation in the WE and SA ethnic groups

WGCNA quantifies both the correlations between individual pairs of genes or probes across a data set, and also the extent to which these probes share the same neighbours (Langfelder and Horvath, 2008). The WGCNA process creates a dendrogram that clusters similarly abundant probes into discrete branches, and subsequent cutting of the dendrogram yields separate co-expression modules, representing putatively co-regulated sets of genes. The first principal component of the expression matrix of each module defines the expression profile of the eigengene for the module, and this can then be correlated with experimental metadata. By allowing phenotypic traits to be associated with relatively small numbers (tens) of module eigengenes, instead of thousands of individual variables (*i.e.* gene probes), WGCNA both alleviates the multiple testing problem associated with standard differential expression analysis and also directly relates experimental traits to gene expression data in an unsupervised way that is agnostic of the experimental design.

WGCNA was used to construct separate signed co-expression networks for the WE and SA ethnic groups as described in the Methods, and Pearson correlations between expression of the module eigengenes and serum 25(OH)D_2_, 25(OH)D_3_, total 25(OH)D and parathyroid hormone (PTH) concentrations were calculated (Fig. 6 and **Supplementary data files 6 - 8**). These results therefore ignore whether the data originates from the vitamins D_2_, D_3_ or placebo treatment groups and focus solely on the relationship between the serum metabolite concentrations and gene expression. A significant negative correlation was observed between 25(OH)D_2_ concentration and expression modules in the WE co-expression network that are enriched for immune-associated functions (midnight blue and pink modules in Fig. 6a). This is consistent with the results from the different analytical approaches (presented in Figs 3 and 4). Similarly, the only module exhibiting a significant positive correlation with 25(OH)D_2_ (and total 25(OH)D concentration), the red expression module, is enriched for GO categories associated with the ribosome and mRNA processing (**Supplementary Data File 6**). No significant correlations with serum 25(OH)D_3_ concentrations were detected in the WE cohort.

**Figure 6.**
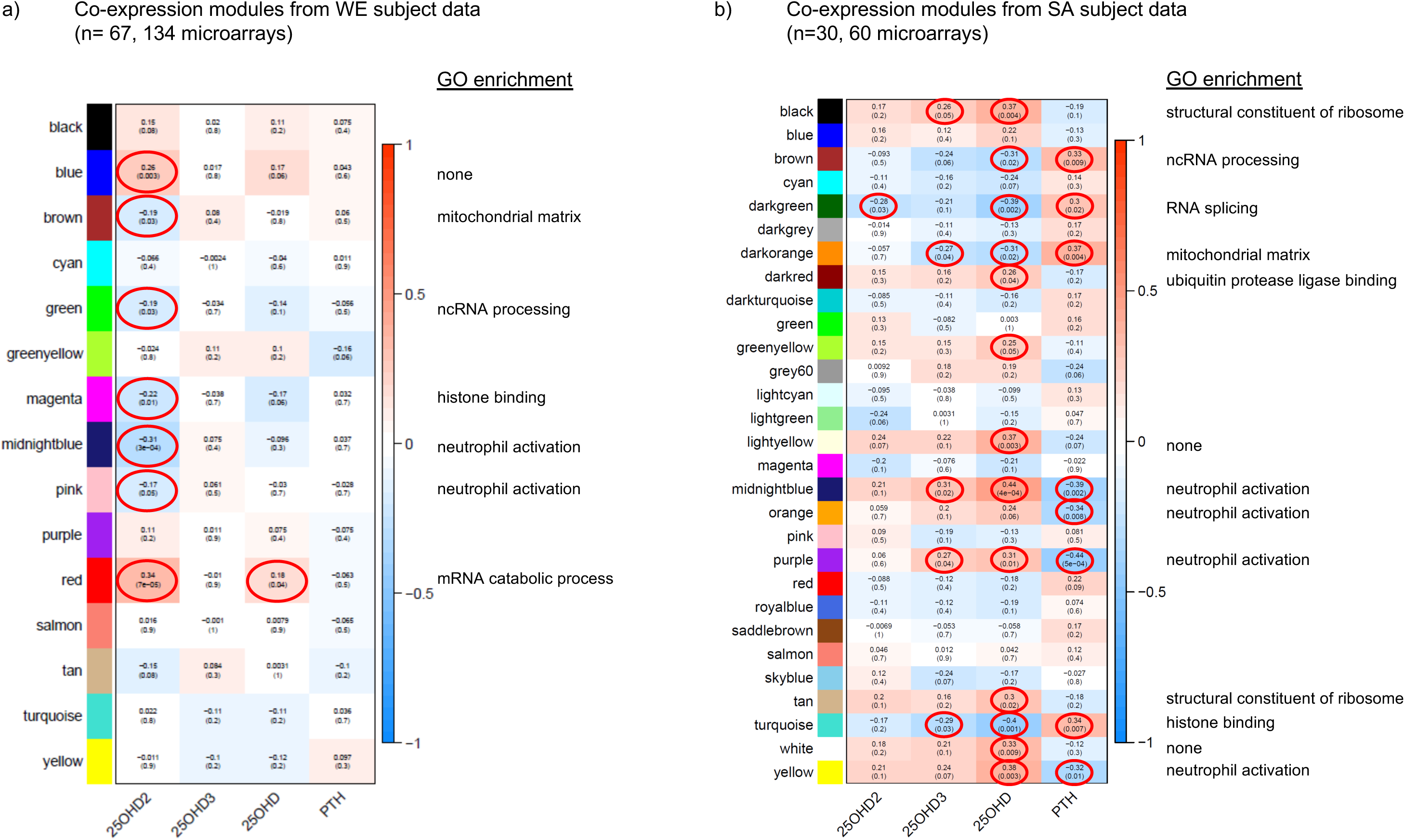
Gene co-expression networks for the WE (a) and SA (b) ethnic groups. For each group of subjects, signed scale-free networks were constructed from the expression data using WGCNA to cluster probes with similar expression characteristics across all samples into discrete modules (colours on the y-axis: colours in (a) and (b) are independent). Module membership for the modules of genes identified in the WGCNA analysis of the data from the South Asian and white European cohorts are detailed in **Supplementary Data File 6**. The colour scale to the right of each panel represents the Pearson correlation (from −1 to +1) between the expression profile for each module’s eigengene and the respective serum 25(OH)D_2_, 25(OH)D_3_, total 25(OH)D and PTH concentrations (x-axis). The Pearson correlation coefficients are also provided in each box, followed in brackets by an adjusted p-value, testing for the significance of each correlation. Statistically significant correlations (adjusted p-value ≤ 0.05) are circled in red. A headline significant GO functional enrichment category (p.adjust ≤ 0.01) for each significant module is shown to the right; GO test results for each module are summarised in **Supplementary Data Files 7 and 8**).

Interestingly, stronger and more numerous correlations were observed in the co-expression network for the SA cohort (Fig. 6b). In agreement with the WE network, an expression module significantly associated with the ribosome (black module in Fig. 6b) showed a general positive correlation with total 25(OH)D concentration whereas a module enriched for histone binding (turquoise) had a significant negative correlation with 25(OH)D concentration. Strikingly however, modules enriched for immune response functions (midnightblue, orange, purple, yellow in Fig. 6b) were consistently positively correlated with total 25(OH)D and usually 25(OH)D_3_, but not 25(OH)D_2_ concentrations in the SA network, and negatively correlated with PTH concentrations. This raises the possibility that vitamin D supplementation may exert different effects on the immune system depending on ethnicity of the individual, may indicate that PTH status has an influence on the outcome and may reflect physiological differences resulting from the low baseline vitamin D status in SA women. In this context we observed that PTH concentrations at the V1 sampling point in the SA cohort were elevated compared with those in the WE cohort (Fig. 1c).

### Gene sets that respond to interferon alpha and gamma show divergent behaviour in the vitamin D_2_ and D_3_ treatment groups of the WE cohort

The molecular signatures database (MSigDB) hallmark gene sets represent coherent gene expression signatures related to well-defined biological states or processes (https://www.gsea-msigdb.org/gsea/msigdb/). Gene set enrichment analysis (GSEA) was performed to identify statistically significant (padjust<=0.05), concordant changes in expression of these hallmark sets between the V3 and V1 sampling times for the placebo, vitamin D_2_, and vitamin D_3_ treatment groups (Fig. 7 and Supplementary Data File 9). Interestingly, in the WE cohort, the gene sets defining the signature responses to interferon alpha and gamma exhibited divergent behaviour in the vitamin D_2_ and D_3_ treatment groups. This is evident as significant up-regulation over the course of the intervention study in the D_3_ group but significant down-regulation in the D_2_ group. This differed from the SA cohort where significant down-regulation was observed in the D_3_ group.

**Figure 7.**
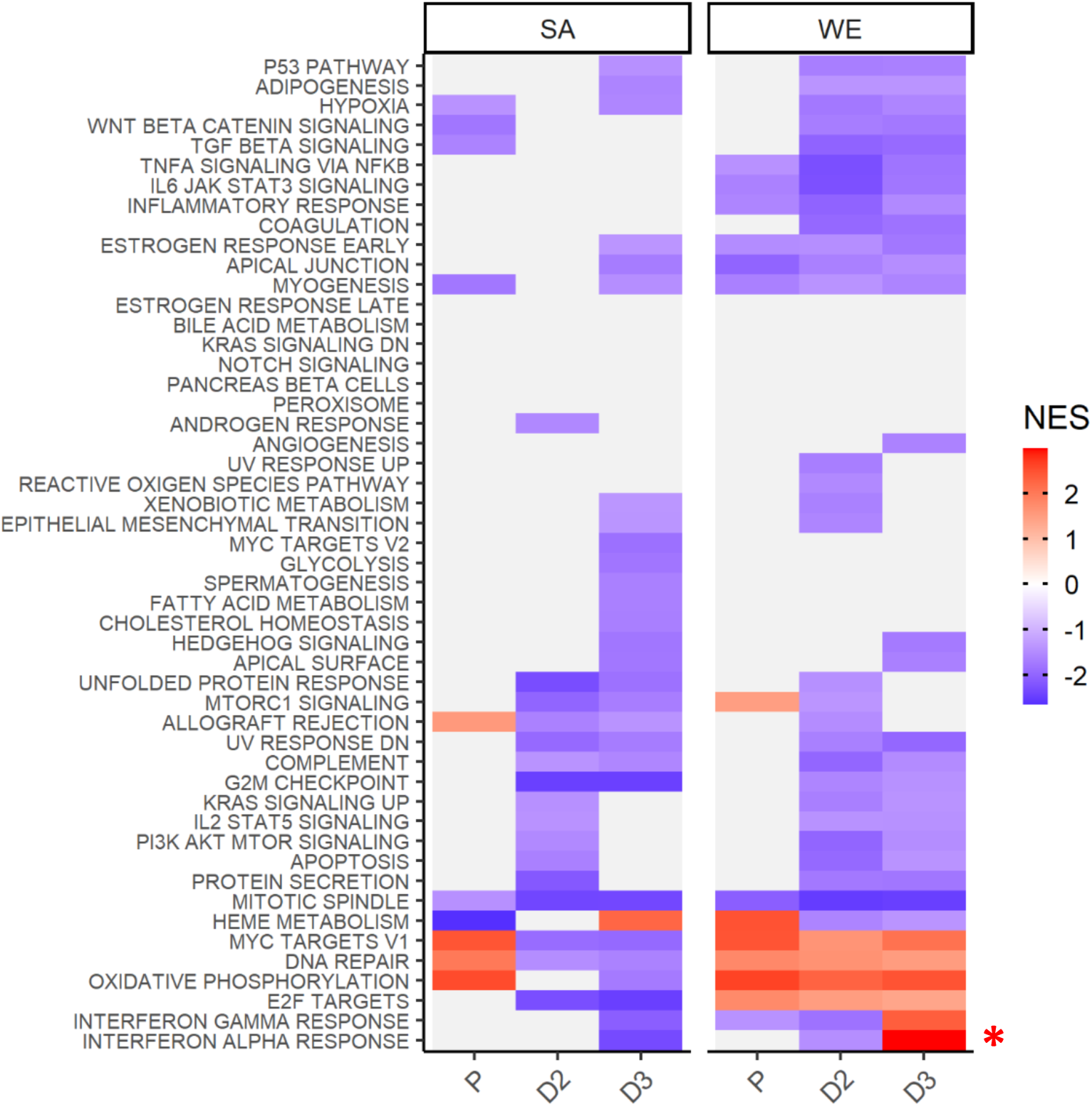
Gene Set Enrichment Analysis using the MSigDB hallmark gene sets indicates divergent behaviour for the interferon alpha and gamma response gene sets following supplementation with vitamin D_2_ and D_3_ in the WE cohort. Coloured tiles in the heatmap correspond to gene sets exhibiting a statistically significant (padjust<=0.05), concordant change between the V3 and V1 sampling times for the placebo (P), vitamin D_2_ (D2) or vitamin D_3_ (D3) treatment groups in the SA or WE cohorts. Grey tiles indicate non-significance. A positive normalised enrichment score (NES) indicates up-regulation of a gene set in V3 relative to V1, and conversely down-regulation is indicated by a negative NES score. Full results are provided in Supplementary Data File 9, and the behavior of the leading edge, core genes accounting for the significant enrichment signals in the interferon alpha and gamma response sets are illustrated in Supplementary Figs. 4-6.

## Discussion

### Vitamin D intake down-regulates expression of many genes including those encoding multiple pathways involved in innate and adaptive immunity

In the present study we shed light on the physiological effects of the two forms of vitamin D commonly used as food-fortificants or vitamin supplements. Using data from the same intervention study, previously we reported that vitamin D_3_ supplementation is more effective than supplementation with vitamin D_2_ in raising serum 25(OH)D concentration (Tripkovic et al., 2017). However, other studies have reported that the two forms of vitamin D are equally effective in raising serum 25(OH)D concentration (Holick et al., 2008, Zittermann et al., 2020). In the present study we examined gene expression changes *in vivo* following supplementation with either vitamin D_2_ or D_3_ (or placebo) in healthy women from the South East of the UK. Here we extend the results reported by Tripkovic et al. (2017) by conducting whole genome transcriptome analysis in 97 of the original cohort of 335 women from two ethnic backgrounds, South Asian (SA) and white European (WE). We characterised gene expression changes in the human blood transcriptome in response to vitamin D_2_ or D_3_ supplementation over a 12-week treatment intervention over the winter months; the randomised groups were provided with physiological (15 µg) daily doses of vitamin D_2_ or vitamin D_3_ fortified foods, or identical foods without added vitamin D (placebo). We found extensive changes in gene expression in all three treatment groups, some of which were unique to the vitamin D_2_-treated or D_3_-treated groups, with the majority exhibiting downregulation of transcription over the 12-week intervention period. Our success in detecting statistically significant differential gene expression in this study stems partly from the longitudinal design where it was possible to evaluate statistical differences in transcript levels within each group (from baseline to 12-week measurement). This approach partly circumvents the problems with inter-individual differences in human studies which make it difficult to detect robust changes between different groups of individuals. Furthermore, the statistical analysis approach examined pairwise differences in gene expression from the V1 and V3 samples, which helps to reduce the impact of inter-individual differences on statistical significance.

This *in vivo* human transcriptome study has demonstrated long-term effects on gene expression. The majority of differentially expressed genes identified in this study were suppressed by vitamin D supplementation, and many of these encoded pathways were involved in immunity. Many of these observed gene expression changes are consistent with vitamin D exerting a modulating effect on the immune system, leading towards a more tolerogenic state, a concept reviewed by (Prietl et al., 2013). The findings from this study are based on an analysis of 97 individuals divided equally between the three treatment groups. Our ability to detect differences in the effects of vitamin D_2_ and vitamin D_3_ may have been negatively impacted by the inclusion of two different ethnic groups among the 97 participants, since the transcriptome results from the two ethnic groups are clearly different. It will be very important to independently replicate this study using a much larger cohort in order to verify, or otherwise, the key findings from this study. From power calculations it is considered that for a whole human microarray-based gene expression study of this nature, and with gene expression changes of the magnitude we observe, at least 400 participants of each ethnic group should be recruited for each treatment, giving a total cohort size of 2,400. The biological interpretation of our findings in the present study should be considered as preliminary in this context, requiring independent verification.

It is difficult to compare the findings of the present study with other reported *in vivo* studies because of the considerable differences in experimental design, including the use of supra-physiological vitamin D doses (up to 2,000 μg single bolus doses), different population types and small sample sizes which make statistical analysis not feasible (e.g. (Hossein-nezhad et al., 2013, Neme et al., 2019)). Furthermore, our study is unique in that it compared gene expression in participants given either of the two commonly used forms of vitamin D, vitamin D_2_ and vitamin D_3_. Intriguing, this revealed that there were considerable differences in the gene expression profiles following each nutritional treatment. For example, excluding genes that were also differentially expressed over the 12-week intervention in the placebo group, only 13% of down-regulated differentially expressed genes were common between the two treatment groups while 28% and 59% were uniquely down-regulated by vitamins D_2_ and D_3_, respectively (Figure 2c).

It is questionable whether it is useful to extrapolate the results from single high dose bolus and *in vitro* studies such as those described by (Koivisto et al., 2020) with this sustained dietary intake *in vivo* study. In the former study 15 ‘vitamin D-responsive’ genes were identified as important mediators of innate and adaptive immunity. Five of these 15 genes are also identified as differentially expressed in our present study (*CD93*, *CEBPB*, *THBD*, *THEMIS2* and *TREM1*). However, in the present study these five genes were *down-regul*ated by vitamin D intake (**Supplementary Data File 2**) whereas they are reported to be up-regulated in the studies reported by (Koivisto et al., 2020). These contrasting results highlight the difficulties with comparing data from different experimental designs. As argued previously, because of the relatively small sample sizes investigated and the small effect sizes, this study has focussed on identification of statistically significant pathway enrichment following sustained vitamin D supplementation, rather than a gene-focussed analysis. Nevertheless, we consider it useful to consider the respective roles of these five specific gene products since their *down*-regulation by vitamin D would make sense from a biological perspective. In our study the gene encoding thymocyte selection-associated family member 2 (*THEMIS2*) is down-regulated after vitamin D_2_ supplementation, the CD93-encoding gene is down-regulated following D_2_ and D_3_ supplementation, while the genes encoding thrombomodulin (*THBD*) and the CCAAT enhancer-binding protein beta (*CEBPB*) are down-regulated following D_3_ supplementation. *THBD* is a key vitamin D target in peripheral blood mononuclear cells (PBMCs) with the function of reducing blood clots while at the same time preventing the pro-inflammatory consequences of NF-kB signalling. The *THBD* and *CD93* genes are next to each other, share the same super-enhancer, and their transcription start site regions respond to 1,25(OH)_2_ D_3_. *CEBPB* encodes a transcription factor important in the regulation of genes involved in immune and inflammatory responses. *TREM1* encodes triggering receptor expressed on myeloid cells which stimulates neutrophil and monocyte-mediated inflammatory responses. It triggers release of pro-inflammatory chemokines and cytokines, as well as increased surface expression of cell activation markers. TREM-1 amplifies inflammatory responses that are triggered by bacterial and fungal infections and is a crucial mediator of septic shock (Bouchon et al., 2001).

Another cellular function that is affected by vitamin D_2_ and D_3_ supplementation is chromatin modification and remodelling. The *EP300* gene is down-regulated by both vitamins, but unaffected in the placebo group. *EP300* encodes a histone acetylase that regulates transcription by chromatin remodelling, in particular histone acetylation of H3 at lysine-27, representing an epigenetic modification which activates transcription (Hatzi et al., 2013). Importantly, EP300 is also a coactivator of VDR.

Functional categories of genes enriched among the upregulated genes, following supplementation with either vitamin D_2_ or D_3,_ include translation, mitochondrial and spliceosome function (Fig. 3b, **Supplementary Data File 4**); statistically enriched biological cellular component terms in these functional categories are ribosomal proteins, components of the mitochondrial respiratory chain, two subunits of the histone H4 and snRNP Sm protein components of the spliceosome assembly. It is established that, in addition to influencing transcription, vitamin D can also influence post-transcriptional events by recruiting co-regulators. In this context it is relevant that components of the spliceosome such as snRNP Sm proteins that mediate both transcriptional control and splicing decisions, leading to alternatively spliced transcripts (Zhou et al., 2015), were upregulated by vitamin D supplementation in this study. These statistically significant gene expression changes were detected as temporal differences *within* each treatment group (differences at baseline (V1) versus 12 weeks after the intervention (V3) *within* single individuals). Differences in gene expression *across* treatment groups were only found in the South Asian group (Fig. 2a, b). The SA treatment group that received supplementation with vitamin D_3_, revealed three genes significantly upregulated and two down-regulated after the 12-week intervention relative to the placebo group (Fig. 2; **Supplementary Fig. 1**). We note that the gene encoding cAMP-responsive element binding protein 1 (*CREB1*) is one of the three upregulated genes. It encodes a transcription factor that binds as a homodimer to the cAMP-responsive element. CREB1 protein is phosphorylated by several protein kinases, and induces transcription of genes in response to hormonal stimulation of the cAMP pathway. The protein kinase A (PKA) pathways are activated by the parathyroid hormone PTH in response to low serum calcium levels to maintain serum calcium homeostasis, primarily by promoting vitamin D 1α-hydroxylation in the kidney. Both PTH and 1,25 (OH)_2_ vitamin D have similar effects in promoting the maturation of osteocytes and opposing the differentiation of osteoblasts into osteocytes (St John et al., 2015, St John et al., 2014). It is relevant to note in this context that all treatment groups in the SA cohort showed elevated serum PTH at the baseline sampling point relative to the WE cohort.

### Vitamins D_2_ and D_3_ do not influence expression of the same genes

Our results indicate that the cellular responses to the two forms of vitamin D supplementation have some commonalities but also show some clear differences (catalogued in **Supplementary Data File 4**). For example, some biological processes such as histone modification and covalent chromatin modification are downregulated following vitamin D_3_ supplementation only, while spliceosomal function are upregulated by vitamin D_2_ only. In light of the finding that vitamins D_2_ and D_3_ influence expression of different genes in the human blood transcriptome, recently we undertook a parallel *in vitro* study with a model rat cell line (Mengozzi et al., 2020). We examined the respective influences of the two physiologically active forms of vitamin D, 1,25(OH)D_3_ and 1,25(OH)D_2_ on differentiation and global gene expression in differentiating rat CG4 oligodendrocyte precursor cells and revealed considerable differences in the influence of the two types of vitamin D on gene expression at 24 h and after 72 h following onset of differentiation. We demonstrated that 1,25(OH)D_3_ and 1,25(OH)D_2_ respectively influenced expression of 1,272 and 574 genes at 24 h following addition of the vitamin to the culture, where many of the changes in expression were specific to one or the other form of the vitamin (Mengozzi et al., 2020). This study provides evidence of the different direct effects of the two active vitamin D metabolites on gene expression *in vitro* in cultured cells and provides some evidence that the changes we observed in the *in vivo* study may reflect the influence of the physiologically active forms of the vitamin, 1,25(OH)D_3_ and 1,25(OH)D_2_.

### Influence of vitamin D on expression of genes encoding immune pathways

In common to both the D_2_ and D_3_ treatment groups, but not the placebo group, we found that many different pathways of the immune system are differentially expressed (largely down-regulated) by vitamin D, consistent with the emerging theme that one physiological role of vitamin D is to suppress, or balance, the activity of the immune system (Prietl et al., 2013, Martens et al., 2020)(Fig 3a, Fig. 4; Supplementary data files 3, 4 and 5). The immunomodulatory effects of vitamin D on both innate and adaptive immunity are well documented (Liu et al., 2006, Hayes and Nashold, 2018, Teles et al., 2013). Vitamin D supplementation has the potential to ameliorate the symptoms of autoimmune diseases such as multiple sclerosis, as supported by several lines of evidence: the role in promoting differentiation of oligodendrocyte progenitor cells and therefore remyelination of CNS neurons (Gomez-Pinedo et al., 2020); suppression of the differentiation of potentially pathogenic T-cells and enhancement of regulatory T-cell differentiation (Wasnik et al., 2020). However, a recent Cochrane review found little benefit of vitamin D in this context (Jagannath et al., 2018).

In this regard, the observation that vitamin D_2_ and D_3_ supplementation in the WE cohort were associated with divergent patterns of expression for interferon alpha (type 1) and interferon gamma (type 2) signature gene sets stands out as particularly interesting. Vitamin D appears to modulate type 1 interferon activity. For example, it enhances the effects of type 1 interferon treatment on mononuclear cells from patients with multiple sclerosis(Feng et al., 2019), which parallels evidence for modest benefits of vitamin D as an adjunct treatment with type 1 interferon in multiple sclerosis patients(Hupperts et al., 2019, Camu et al., 2019). Vitamin D may also help suppress symptoms in autoimmune diseases such as systemic lupus erythematosus(Zheng et al., 2019), which are associated with chronic over-activity of interferon signalling and tentatively designated interferonopathies. Moreover, type 1 interferons play a critical role in defence against viral infections. Basal expression of type 1 interferon-stimulated genes, in the absence of infection, is key to priming a rapid and effective response to viral infection when it occurs(Mostafavi et al., 2016). There is currently intense interest in vitamin D as both a potential prophylactic and a therapeutic agent for treatment of SARS-CoV-2 infection. One of the proposed modes of action of vitamin D is modulation of interferon activity(Gauzzi and Fantuzzi, 2020); in this context our observation that vitamin D_3_ (but not vitamin D_2_) enhances the expression of genes involved in the interferon alpha response, is highly relevant to susceptibility to viral infection. Indeed, a recent genome-wide association study (GWAS) in 2,244 critically ill Covid-19 patients identified genetic variants leading to reduced interferon type 1 signalling that are associated with severe Covid-19 disease(Pairo-Castineira et al., 2020).

We have also found differences in gene expression response to vitamin D according to ethnicity (with the caveat that the sample size of the SA group was smaller than the WE group). Ethnic differences in response are suggested from the Weighted Gene Correlation Network Analysis (WGCNA) and the Gene Set Enrichment Analysis (GSEA), where the response to vitamin D_3_ intake appeared to have the opposite effect on the type 1 and 2 interferon pathways in the SA group compared with the WE group (Fig. 7). However, direct correlation between the stimulation of immune responses, other than the interferon pathways, with an increase in serum 25(OH)D_3_ concentration was evident in the SA group only. This is the opposite of that observed in the WE group where the effect of vitamin D supplementation was to suppress several immune pathways.

## Conclusions

The transcriptome results from this study differ from those of previous *in vivo* studies which used supra-physiological doses of vitamin D over a short time-frame, or used *in vitro* cultured cells. Indeed, where there is overlap in the identity of the differentially expressed genes identified in the respective studies vitamin D appears to exert the opposite effect relative to our longer-term repeated physiological dose study. Short-term responses to large boluses are relatively simple to do, and less prone to noise, but their effect on gene expression does not seem to be consistent with that observed with longer term repeated physiological dose studies like the present study and therefore may not represent the real-world effects of repeated dietary intake of vitamin D. Thus, despite the challenges and limitations of doing longer term *in vivo* human studies, that are complicated by seasonal variation in gene expression, inter-individual human variation, subtle physiological changes in gene expression, more ‘noise’ through intake of other vitamin D sources during the treatment intervention, our work suggests it may be important to conduct more such longer term studies since the results from the shorter term interventions may not be representative of the influence of vitamin D on normal physiological gene expression. One potential criticism of whole blood transcriptome studies is that the results may not represent the gene expression changes that occur in other tissues as a result of vitamin D supplementation. However, we have shown, in comparable whole blood transcriptome studies of human sleep physiology, that the changes in the human blood transcriptome correlate with the changes that are observed in other tissues and organs in comparable mammalian animal studies, giving us some confidence in the physiological relevance of the changes identified in this vitamin D transcriptome study (Archer et al., 2014, Moller-Levet et al., 2013).

The transcriptome results differ considerably between the two different ethnic groups. While some of the differences may be attributable to differences in sample sizes studied between the two groups it is also possible that some of the differences represent genuine differences in the respective physiological responses of the two ethnic groups. Alternatively, the differences may reflect the starting physiological status of the SA participant group, who were found to have considerably lower baseline vitamin D concentrations (and higher PTH concentrations) than the WE group. The importance of clarifying these different responses is given particular impetus by the emerging evidence for the interplay between ethnicity, skin tone, vitamin D status and susceptibility to viral infection, Covid-19 being of particular current relevance here (Lanham-New et al., 2020, Kohlmeier, 2020, Kmietowicz, 2020, Weir et al., 2020). The transcriptomic data presented in this study might provide a useful context for further studies aimed at understanding the role of vitamin D in influencing the immune response to SARS-CoV-2 infection, particularly in relation to severe Covid-19. It is notable that the recent GenOMICC study of severe Covid-19 disease highlighted a link between severe Covid-19 and reduced type 1 interferon signalling (Pairo-Castineira et al., 2020). Our finding that vitamin D_3_ appears to stimulate type 1 interferon signalling could be relevant in the context of its use as a prophylactic treatment.

In summary our results indicate that whilst the two forms of vitamin D (D_2_ and D_3_) exert some overlapping roles in human physiology, each form may elicit different molecular responses, findings which warrant a more detailed consideration of the system-wide effects of these two forms vitamin D. Clearly, this type of study requires replication, using a much larger independent cohort, with balanced representation from different ethnic groups. This is perhaps of particular importance to at-risk ethnic groups, including black and South Asian populations who reside in northern latitudes. The studies would need to be time course-based (temporal) to track gene expression changes *within* individuals from baseline activity to defined sampling times and would need in-built control to account for seasonal gene expression changes. They would need to be designed specifically to answer whether different ethnicity or different vitamin D baseline levels give rise to different responses to vitamin D supplementation.

Since some pathways appear to be regulated specifically by vitamin D_3,_ or in some cases, in opposing directions by vitamin D_3_ and D_2_, future studies should investigate whether vitamin D_2_ supplementation might counteract some of the benefits of vitamin D_3_ on human health. This possibility is prompted by the findings from this cohort that the circulating concentration of 25(OH)D_3_ within vitamin D_2_-treated participants was significantly lower after the 12-week intervention than in the placebo group who received no vitamin D supplements – suggesting that the former might be depleted by the latter. The results from this study suggest that guidelines on food fortification and supplementation with specific forms of vitamin D may need revisiting.

## Materials and Methods

### Blood Transcriptome analysis

The recruitment of individuals as part of the BBSRC D2-D3 Study was described previously (Tripkovic et al., 2017). This study received ethical approval from the South-East Coast (Surrey) National Health Service Research Ethics Committee (11/LO/0708) and the University of Surrey Ethics Committee (EC/2011/97/FHMS). All of the participants gave written informed consent in agreement with the Helsinki Declaration before commencing study activities. Briefly, 335 women of both South Asian (SA) and white European (WE) descent were randomised to one of three intervention groups for 12-weeks and provided with daily doses of vitamin D within fortified foods: placebo, 15 μg/d vitamin D2 or 15 μg/d vitamin D3. Of the 335 participants, 97 were selected for transcriptome analysis (32 were WE, 32 were A and 33 were selected at random from both ethnic groups to form a balanced placebo group). High responders (>50% increase in 25OHD concentrations) and low responders (<50% increase in 25OHD concentrations) were included within both vitamin D_2_ and D_3_ supplementation groups to maximize the chance of observing differences in gene expression among subjects. The selected time points for transcriptome analysis were V1 representing the baseline and V3, 12 weeks after treatment commenced.

Whole peripheral blood (2.5 ml) was collected using PAXgene Blood RNA tubes (Becton Dickinson). PAXgene Blood RNA Tubes were inverted ten times immediately after drawing blood, stored upright at 15-25°C for 24 hours, followed by a −20°C freezer for 24 hours and then into a −80°C freezer for long-term storage.

Transcriptomic analysis was conducted essentially as described in previous studies (Archer et al., 2014, Moller-Levet et al., 2013). Total RNA was isolated using the PAXgene Blood RNA Kit (Qiagen) following the manufacturers recommendations. cRNA was synthesised and fluorescently labelled with Cy3-CTP from 200 ng of total RNA using the Low Input QuickAmp Labelling Kit, One Color (Agilent Technology). Labelled cRNA was hybridised on a Sure Print G3 Human Gene Expression 8 x 60K v2 microarray slide (Agilent Technologies). Standard manufacturer’s instructions for one colour gene-expression analysis were followed for labelling, hybridisation and washing steps. Extracted RNA was quantified using NanoDrop ND2000 spectrophotometer (Thermo Scientific). RNA quality and integrity was evaluated using either the Bioanalyzer 2100 or the TapeStation 4200 (Agilent Technologies). Only RNA samples with an RNA Integrity Number (RIN) of >7.0 were subjected to DNA microarray analysis. Microarrays were hybridised at 65°C for 17 hours in an Agilent Hybridization Oven on a rotisserie at 10 rpm. The washed microarrays were scanned using an Agilent Microarray Scanner with a resolution of 2 μm.

### Transcriptome data processing and differential expression analysis

Raw scanned microarray images were processed using Agilent Feature Extraction software (v11.5.1.1) with the Agilent 039494_D_F_20140326_human_8×60K_v2 grid, and then imported into R for normalization and analysis using the LIMMA package (Ritchie et al., 2015). Microarray data were background-corrected using the ‘normexp’ method (with an offset of 50) and quantile normalised, producing expression values in the log base 2 scale. The processed data were then filtered to remove probes exhibiting low signals across the arrays, retaining non-control probes that are at least 10% brighter than negative control probe signals on at least 41 arrays (∼20% of the arrays in the analysis). Data from identical replicate probes were then averaged to produce expression values at the unique probe level. Initial data exploration identified one sample (participant 0017, V3 time point) with array data that was a notable outlier from the group and therefore both the V1 and V3 microarrays for this subject were excluded from all subsequent analysis, reprocessing the data as above in their absence before proceeding.

Tests for differential expression were performed using LIMMA, applying appropriate linear model designs to identify: (i) significant differences in the transcriptional responses occurring across the 12-week V1 to V3 period of the study between the treatment and placebo groups for each ethnic cohort (example contrasts tested, in the format ethnicity_treatment_time: (WE_D2_V3 - WE_D2_V1) - (WE_P_V3-WE_P_V1) = 0, and (WE_D3_V3 - WE_D3_V1) - (WE_P_V3 - WE_P_V1) = 0); and ii) to determine significant changes for each ethnic cohort within each treatment group between the V3 and V1 sampling points, blocking on subject identity (example contrasts tested: WE_D2_V3 - WE_D2_V1 = 0, WE_D3_V3 - WE_D3_V1 = 0). Blocking on subject identity was used to control for inter-subject variability. Significance p-values were corrected for multiplicity using the Benjamini and Hochberg method, obtaining adjusted p-values (adj.P.Val). Normalised data for all participants (V3 – V1) was assessed by principal component analysis to screen for any batch effects (**Supplementary Fig. 3**).

### Functional enrichment and network analysis

Functional enrichment analysis of lists of genes of interest possessing valid ENTREZ gene identifiers was performed using the R package clusterProfiler (Yu et al., 2012). The software produces adjusted p-values (p.adjust) using the Benjamini and Hochberg correction method. Construction and analysis of protein-protein interaction networks from sets of genes was undertaken in Cytoscape (Shannon et al., 2003) using the STRING plugin (Szklarczyk et al., 2017a). Cytoscape was also used to construct and visualise commonality in the functional categories identified as being significantly enriched in the genes responding to the experimental treatments. To visualise categories identified from the D_2_ or D_3_ treatment groups but not from the placebo (e.g. Figs. 4 and 5) significantly enriched categories (p.adjust < 0.01) from all groups were imported such that treatment group nodes (D_2_, D_3_ and placebo (P)) are linked to functional category nodes by edges assigned the corresponding p.adjust values. All nodes with an edge connection to the placebo treatment node were then removed and the resulting networks further filtered to retain only those nodes with at least one edge connection with p.adjust<=0.001.

Weighted gene co-expression network analysis was performed in R using the WGCNA (Langfelder and Horvath, 2008) and CoExpNets (Botia, 2019) packages. A normalised expression data matrix generated by filtering to retain probes with signals more markedly above background (30% brighter than negative control probe signals on at least 41 arrays) was used as input, consisting of 12,169 unique probes. Signed scale-free networks were constructed separately for the data for the SA and WE ethnic groups using 50 iterations of the k-means clustering option in the CoExpNets ‘getDownstreamNetwork’ function to refine the clustering process (**Supplementary Data File 6**). Pearson correlations between cluster module eigengenes and metadata for serum 25(OH)D_2_, 25(OH)D_3_, total 25(OH)D and PTH concentrations were calculated using pairwise complete observations.

### Gene set enrichment analysis (GSEA)

GSEA was performed using the R package clusterProfiler (Yu et al., 2012), applying the default parameters which implement the fgsea algorithm and correct for multiple testing using the Benjamini and Hochberg method. Gene sets were obtained from MSigDB via the msigdbr package (Dolgalev, 2020). Ranked gene lists for interrogation were derived from LIMMA analysis of the unfiltered microarray data, pre-ranking genes according to their t-statistic.

## Data Availability

All details of participants and their metadata are archived in a previous publication (https://doi.org/10.3945/ajcn.116.138693) and the pertinent data for this publication also forms supplementary data files linked with this submission. The full transcriptome data sets have been deposited with the international repository ArrayExpress and allocated accession number: E-MTAB-8600

http://www.ebi.ac.uk/arrayexpress

## Data availability

All microarray data are available in the ArrayExpress database (http://www.ebi.ac.uk/arrayexpress) under accession number E-MTAB-8600.

## Acknowledgements

This work was funded by BBSRC (UK) DRINC award BB/I006192/1 to SLN and CPS, by a University of Brighton Innovation Seed Fund Award (to CPS). The authors would like to particularly thank Michael Chowen CBE DL and Maureen Chowen for their generous philanthropic donation (to CPS).

## Supplementary Figures and Supplementary Data Files

**Supplementary Figure 1.**

Summary of the probes identified as being significantly different in the ‘difference-in-difference’ analysis of the SA cohort (comparison [SA D3 V3 v V1] v [SA P V3 v V1] in Fig. 2a). (A) Differential expression test results; (B) Probe signal abundance (log_2_) distribution across the experimental treatments in the SA cohort.

**Supplementary Figure 2.**

Seasonal gene expression as a feature of the 12-week study (P = placebo, D2 = vitamin D_2_, D3 = vitamin D_3_). (a) Summary of the timing of the V1 samples taken in the study (for each individual, V3 samples were taken 12 weeks after the V1 sample); (b) Significant over-representation of the seasonal genes identified by Dopico et al (2015) *Nat Commun*, **6**, 7000 (BABYDIET dataset) in the genes identified as significantly changing in the white European (WE) placebo, D_2_ and D_3_ treatment groups.

**Supplementary Figure 3.**

Principal component analysis of the transcriptome data for the 97 subjects passing quality control. Data for samples from visits V1 and V3 are processed as the difference between V3 and V1 (V3 minus V1) for each subject. The distribution is relatively random.

**Supplementary Figure 4.**

The MSigDB Hallmark gene sets “Interferon alpha response” and “Interferon gamma response” show statistically significant, concordant up-regulation between the V3 and V1 sampling times following supplementation with vitamin D_3_ in the WE cohort (adjusted p-values 9.36e-15 and 1.59e-11, respectively). a) Leading edge, core genes accounting for the enrichment signals in the gene set enrichment analysis of the D_3_ data. b) Heatmap showing the log2 fold change between V3 and V1 for each gene identified in a), for all the treatment groups in both the SA and WE cohorts (P = placebo, D2 = vitamin D_2_, D3 = vitamin D_3_). Gene membership of the Hallmark interferon alpha and gamma response gene sets is indicated to the right. Genes shared between both sets are labelled “alpha & gamma”. Gene PARP14 and ISG20 belong to both sets, but are leading edge genes only for the alpha response.

**Supplementary Figure 5.**

The MSigDB Hallmark gene sets “Interferon alpha response” and “Interferon gamma response” show statistically significant, concordant down-regulation between the V3 and V1 sampling times following supplementation with vitamin D_2_ in the WE cohort (adjusted p-values 4.22e-02 and 1.61e-04 respectively). a) Leading edge, core genes accounting for the enrichment signals in the gene set enrichment analysis. b) Heatmap showing the log2 fold change between V3 and V1 for each gene identified in a), for all the treatment groups in both the SA and WE cohorts (P = placebo, D2 = vitamin D_2_, D3 = vitamin D_3_). Gene membership of the Hallmark interferon alpha and gamma response gene sets is indicated to the right. Genes shared between both sets are labelled “alpha & gamma”. Gene PARP14 belongs to both sets, but is a leading edge gene only for the alpha response.

**Supplementary Figure 6.**

The MSigDB Hallmark gene sets “Interferon alpha response” and “Interferon gamma response” show statistically significant, concordant down-regulation between the V3 and V1 sampling times following supplementation with vitamin D_3_ in the SA cohort (adjusted p-values 1.34e-09 and 8.45e-09 respectively). (a) Leading edge, core genes accounting for the enrichment signals in the gene set enrichment analysis. (b) Heatmap showing the log_2_ fold change between V3 and V1 for each gene identified in (a), for all the treatment groups in both the SA and WE cohorts (P = placebo, D2 = vitamin D_2_, D3 = vitamin D_3_). Gene membership of the Hallmark interferon alpha and gamma response gene sets is indicated to the right. Genes shared between both sets are labelled “alpha & gamma”. Gene PSMB8 belongs to both sets, but is a leading edge gene only for the alpha response.

## Supplementary Data Files

**Supplementary Data File 1**

Metadata on serum concentrations of 25(OH)D_2_, 25(OH)D_3_, total 25(OH)D, PTH and calcium (albumin-adjusted) for the 97 subjects subjected to transcriptomic analysis (selected from the cohort described by Tripkovic et al. 2017).

**Supplementary Data File 2** [Note: data file is 9.1 MB.]

Significant differences in the transcriptional responses occurring across the 12-week V1 to V3 period of the study *between* the vitamin D_2_, vitamin D_3_ and placebo treatment groups for each ethnic cohort, and significant changes *within* each group between the V3 and V1 sampling points (see Fig. 2).

**Supplementary Data File 3**

Functional analysis of the genes represented by the probes specifically repressed by D_2_ and D_3_ in the white European cohort (blue data points in Fig. 2d) or induced by D_2_ and D_3_ in the white European cohort (red data points in Fig. 2d).

**Supplementary Data File 4**

Comparative functional enrichment analysis of all significant changes in each treatment group in the white European cohort (Fig. 2c; and see Figs 4 and 5).

**Supplementary Data File 5**

Networks illustrating all the functional categories significantly enriched (p.adjust < 0.01) in analysis of the gene products represented by the probes significantly up- or down-regulated (adj.P.Val < 0.05) in the white European cohort treatment groups (from data presented in **Supplementary Data File 4**).

**Supplementary Data File 6**

Module membership for the modules of genes identified in the WGCNA analysis of the data from the South Asian and white European cohorts, as presented in **Fig. 6**.

**Supplementary Data File 7**

Gene ontology functional enrichment analysis results for the modules of genes identified in the WGCNA analysis of the data from the white European cohort.

**Supplementary Data File 8**

Gene ontology functional enrichment analysis results for the modules of genes identified in the WGCNA analysis of the data from the South Asian cohort.

**Supplementary Data File 9**

Gene Set Enrichment Analysis results for the analysis reported in Fig. 7.

