## Supplementary Figures for "Vitamins D_2_ and D_3_ have overlapping but different effects on human gene expression revealed through analysis of blood transcriptomes in a randomised double-blind placebo-controlled food-fortification trial"

### Slide 1
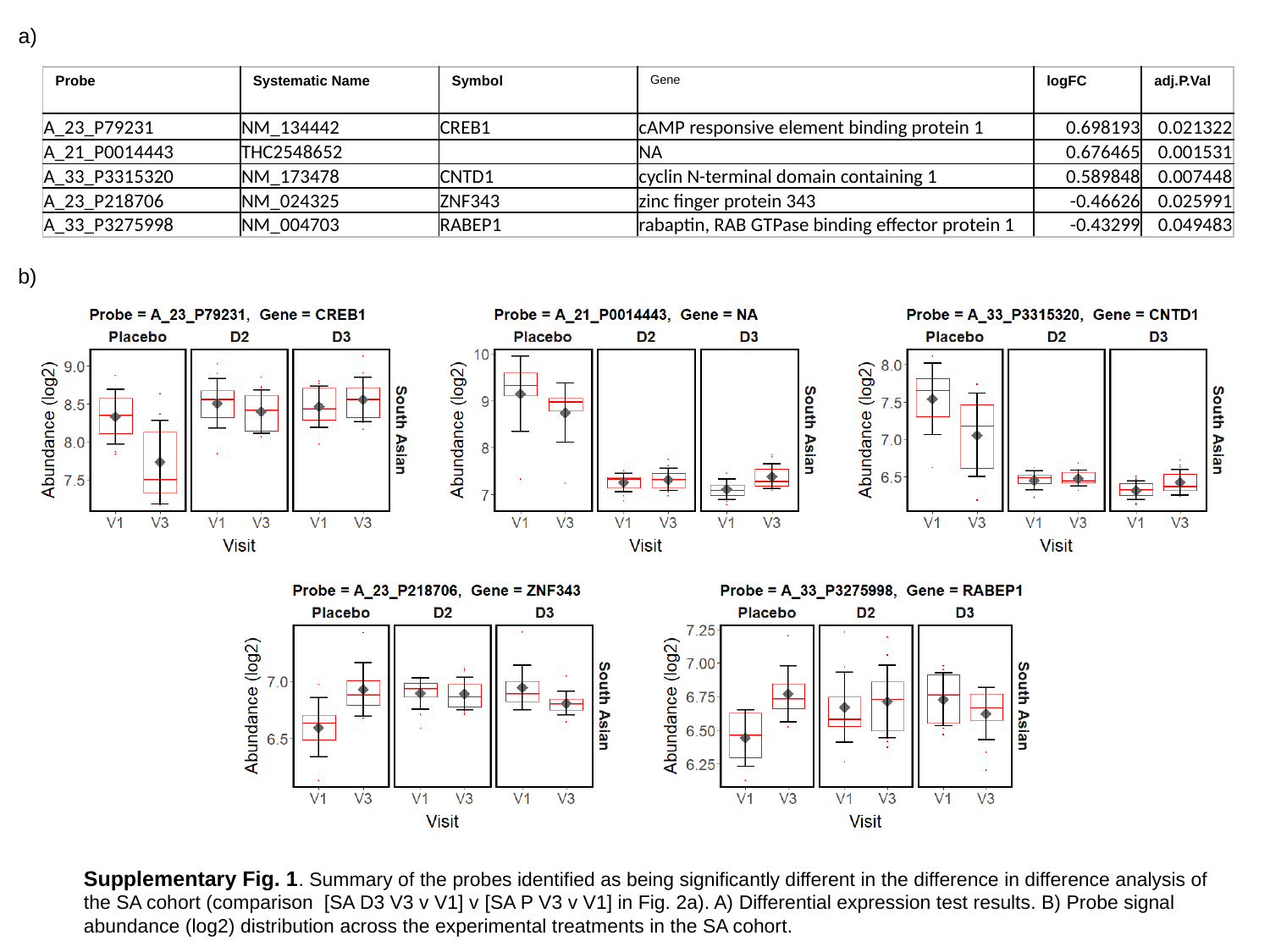

a)
| Probe | Systematic Name | Symbol | Gene | logFC | adj.P.Val |
| --- | --- | --- | --- | --- | --- |
| A\_23\_P79231 | NM\_134442 | CREB1 | cAMP responsive element binding protein 1 | 0.698193 | 0.021322 |
| A\_21\_P0014443 | THC2548652 | | NA | 0.676465 | 0.001531 |
| A\_33\_P3315320 | NM\_173478 | CNTD1 | cyclin N-terminal domain containing 1 | 0.589848 | 0.007448 |
| A\_23\_P218706 | NM\_024325 | ZNF343 | zinc finger protein 343 | -0.46626 | 0.025991 |
| A\_33\_P3275998 | NM\_004703 | RABEP1 | rabaptin, RAB GTPase binding effector protein 1 | -0.43299 | 0.049483 |
b)
Supplementary Fig. 1. Summary of the probes identified as being significantly different in the difference in difference analysis of the SA cohort (comparison [SA D3 V3 v V1] v [SA P V3 v V1] in Fig. 2a). A) Differential expression test results. B) Probe signal abundance (log2) distribution across the experimental treatments in the SA cohort.

### Slide 2
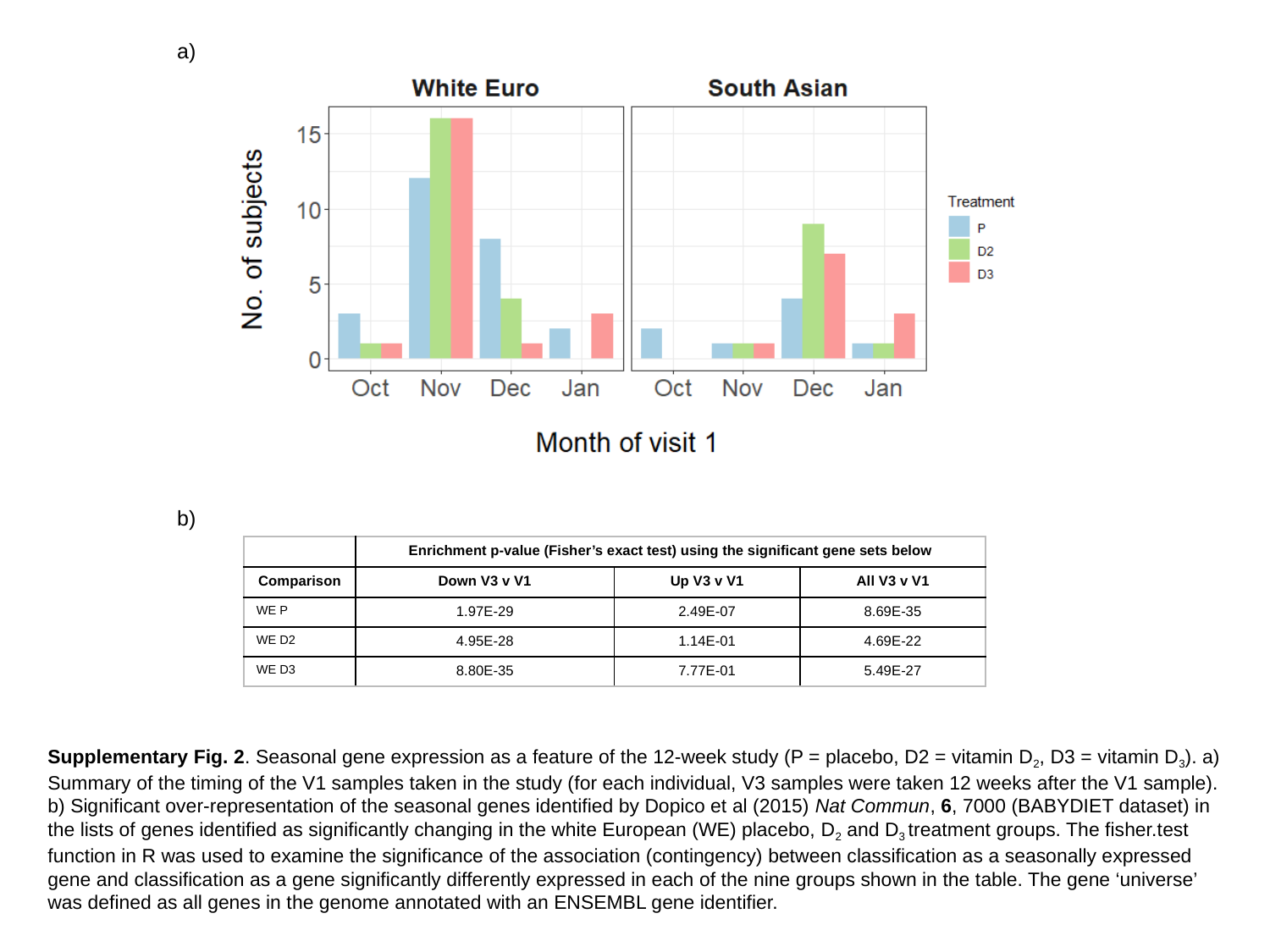

a)
b)
| | Enrichment p-value (Fisher’s exact test) using the significant gene sets below | | |
| --- | --- | --- | --- |
| Comparison | Down V3 v V1 | Up V3 v V1 | All V3 v V1 |
| WE P | 1.97E-29 | 2.49E-07 | 8.69E-35 |
| WE D2 | 4.95E-28 | 1.14E-01 | 4.69E-22 |
| WE D3 | 8.80E-35 | 7.77E-01 | 5.49E-27 |
Supplementary Fig. 2. Seasonal gene expression as a feature of the 12-week study (P = placebo, D2 = vitamin D_2_, D3 = vitamin D_3_). a) Summary of the timing of the V1 samples taken in the study (for each individual, V3 samples were taken 12 weeks after the V1 sample). b) Significant over-representation of the seasonal genes identified by Dopico et al (2015) Nat Commun, 6, 7000 (BABYDIET dataset) in the lists of genes identified as significantly changing in the white European (WE) placebo, D_2_ and D_3_ treatment groups. The fisher.test function in R was used to examine the significance of the association (contingency) between classification as a seasonally expressed gene and classification as a gene significantly differently expressed in each of the nine groups shown in the table. The gene ‘universe’ was defined as all genes in the genome annotated with an ENSEMBL gene identifier.

### Slide 3
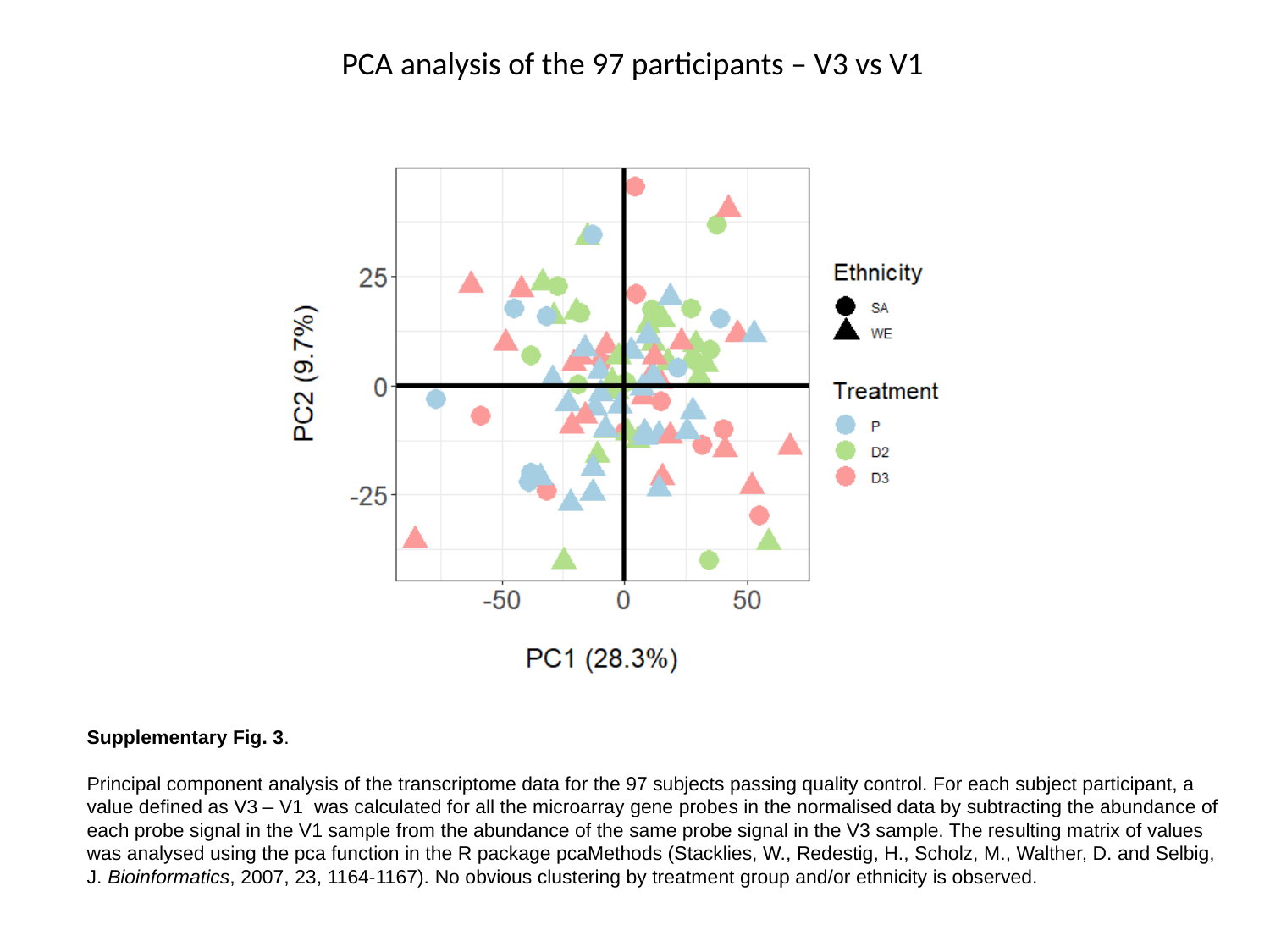

PCA analysis of the 97 participants – V3 vs V1
Supplementary Fig. 3.
Principal component analysis of the transcriptome data for the 97 subjects passing quality control. For each subject participant, a value defined as V3 – V1 was calculated for all the microarray gene probes in the normalised data by subtracting the abundance of each probe signal in the V1 sample from the abundance of the same probe signal in the V3 sample. The resulting matrix of values was analysed using the pca function in the R package pcaMethods (Stacklies, W., Redestig, H., Scholz, M., Walther, D. and Selbig, J. Bioinformatics, 2007, 23, 1164-1167). No obvious clustering by treatment group and/or ethnicity is observed.

### Slide 4
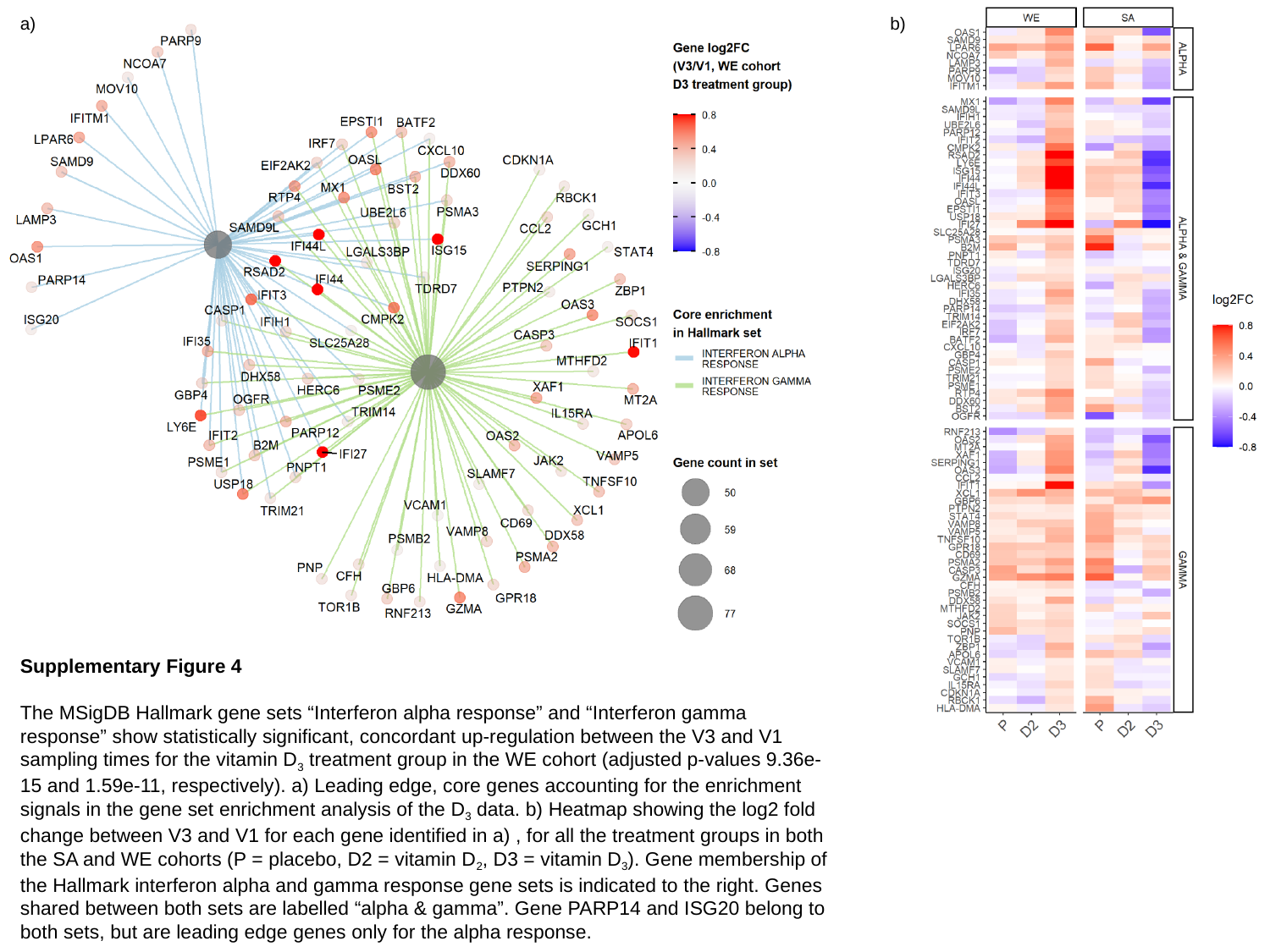

a)
b)
Supplementary Figure 4
The MSigDB Hallmark gene sets “Interferon alpha response” and “Interferon gamma response” show statistically significant, concordant up-regulation between the V3 and V1 sampling times for the vitamin D_3_ treatment group in the WE cohort (adjusted p-values 9.36e-15 and 1.59e-11, respectively). a) Leading edge, core genes accounting for the enrichment signals in the gene set enrichment analysis of the D_3_ data. b) Heatmap showing the log2 fold change between V3 and V1 for each gene identified in a) , for all the treatment groups in both the SA and WE cohorts (P = placebo, D2 = vitamin D_2_, D3 = vitamin D_3_). Gene membership of the Hallmark interferon alpha and gamma response gene sets is indicated to the right. Genes shared between both sets are labelled “alpha & gamma”. Gene PARP14 and ISG20 belong to both sets, but are leading edge genes only for the alpha response.

### Slide 5
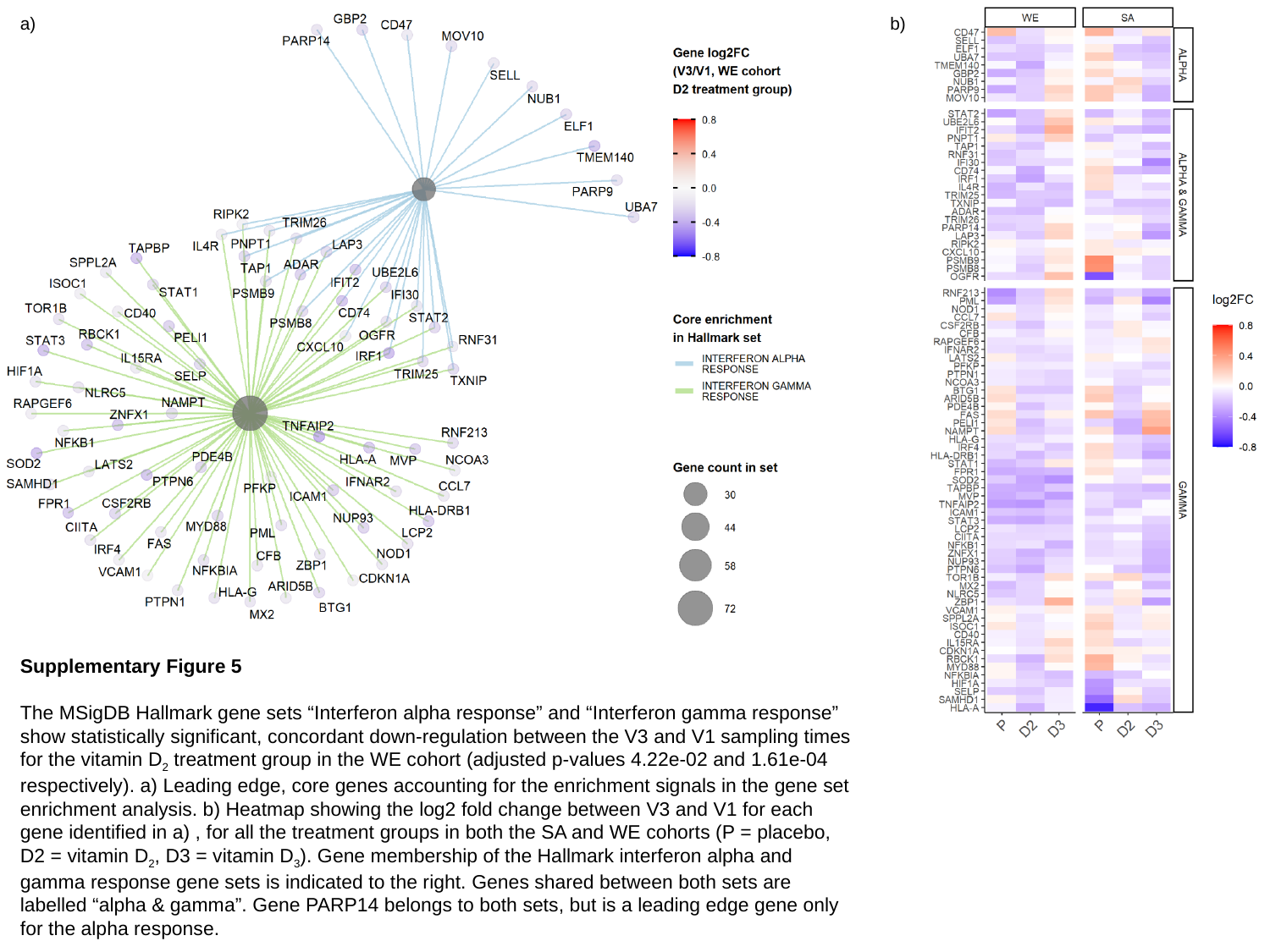

a)
b)
Supplementary Figure 5
The MSigDB Hallmark gene sets “Interferon alpha response” and “Interferon gamma response” show statistically significant, concordant down-regulation between the V3 and V1 sampling times for the vitamin D_2_ treatment group in the WE cohort (adjusted p-values 4.22e-02 and 1.61e-04 respectively). a) Leading edge, core genes accounting for the enrichment signals in the gene set enrichment analysis. b) Heatmap showing the log2 fold change between V3 and V1 for each gene identified in a) , for all the treatment groups in both the SA and WE cohorts (P = placebo, D2 = vitamin D_2_, D3 = vitamin D_3_). Gene membership of the Hallmark interferon alpha and gamma response gene sets is indicated to the right. Genes shared between both sets are labelled “alpha & gamma”. Gene PARP14 belongs to both sets, but is a leading edge gene only for the alpha response.

### Slide 6
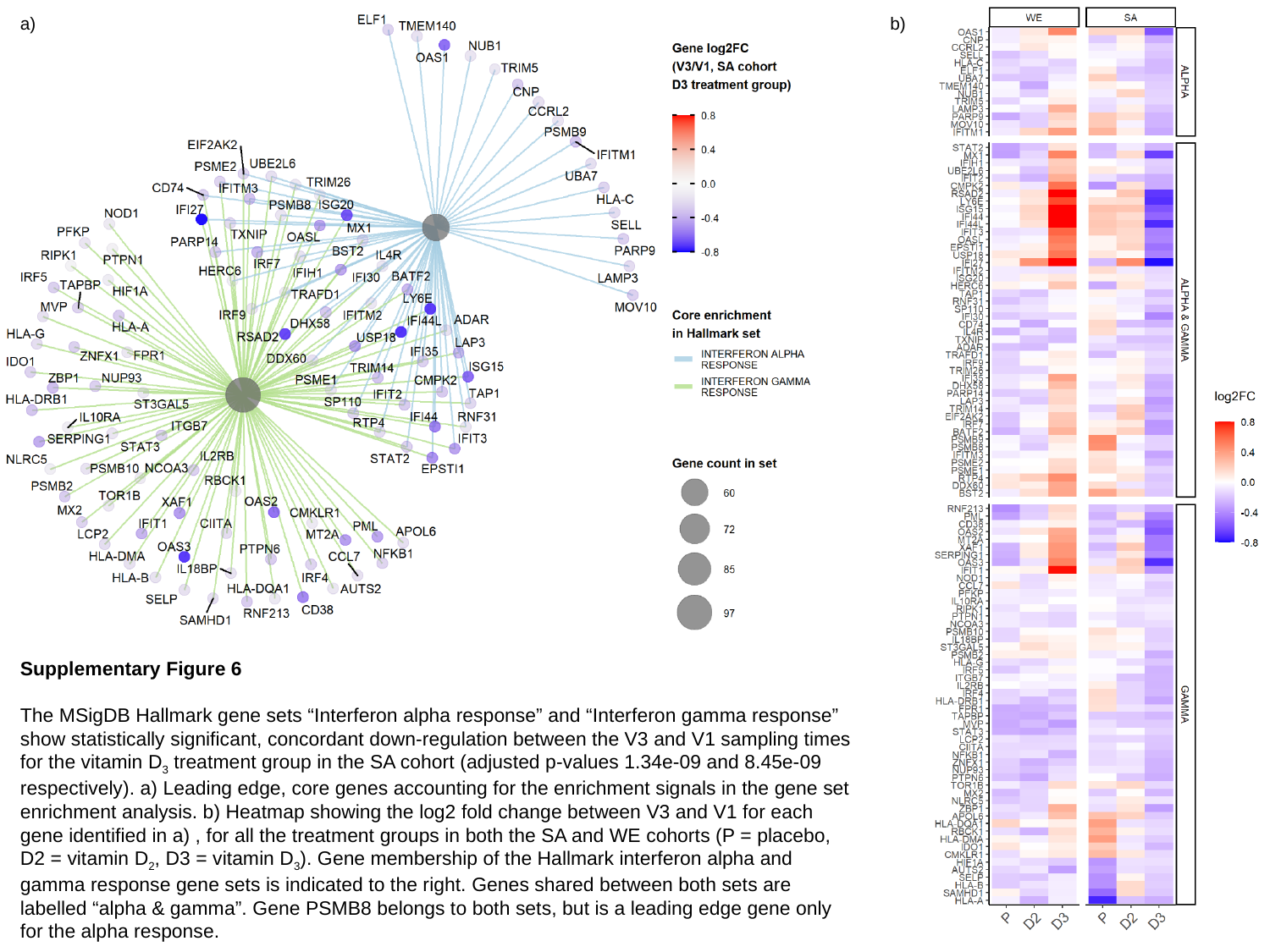

a)
b)
Supplementary Figure 6
The MSigDB Hallmark gene sets “Interferon alpha response” and “Interferon gamma response” show statistically significant, concordant down-regulation between the V3 and V1 sampling times for the vitamin D_3_ treatment group in the SA cohort (adjusted p-values 1.34e-09 and 8.45e-09 respectively). a) Leading edge, core genes accounting for the enrichment signals in the gene set enrichment analysis. b) Heatmap showing the log2 fold change between V3 and V1 for each gene identified in a) , for all the treatment groups in both the SA and WE cohorts (P = placebo, D2 = vitamin D_2_, D3 = vitamin D_3_). Gene membership of the Hallmark interferon alpha and gamma response gene sets is indicated to the right. Genes shared between both sets are labelled “alpha & gamma”. Gene PSMB8 belongs to both sets, but is a leading edge gene only for the alpha response.
