## Supplemental data file 5 for "Vitamins D_2_ and D_3_ have overlapping but different effects on human gene expression revealed through analysis of blood transcriptomes in a randomised double-blind placebo-controlled food-fortification trial"

**Supplementary Data File 5**

Networks illustrating the functional categories significantly enriched in the gene products represented by the probes significantly up-regulated or down-regulated (adj.P.Val <= 0.05) in the $D_2$ and $D_3$ treatment groups of the WE cohort (but not Placebo group).

Gene products represented by the significantly up- or down-regulated probes in the comparisons WE D2 V3 v V1, WE D3 V3 v V1 and WE P V3 v V1 from Fig. 2(a), and possessing ENTREZ identifiers, were subjected separately to functional enrichment analysis using compareCluster and processed In Cytoscape (see legend to **Fig. 4**). The complete networks for each differentially expressed group of genes are shown on pages 3 – 12.

Example of naming convention used below: **cc.WE.PD2D3.down.GOBP** are the down-regulated genes for each of the P, D2 and D3 categories, analysed by gene ontology enrichment using biological process categories.

**Note**: *the images are vector graphics and you can therefore zoom in to read the different functional groups*


| Page | |
|---|---|
| 3 | cc.WE.PD2D3.down.GOBP.pdf |
| 4 | cc.WE.PD2D3.down.GOCC.pdf |
| 5 | cc.WE.PD2D3.down.GOMF.pdf |
| 6 | cc.WE.PD2D3.down.KEGG.pdf |
| 7 | cc.WE.PD2D3.down.PWAY.pdf |
| 8 | cc.WE.PD2D3.up.GOBP.pdf |
| 9 | cc.WE.PD2D3.up.GOCC.pdf |
| 10 | cc.WE.PD2D3.up.GOMF.pdf |
| 11 | cc.WE.PD2D3.up.KEGG.pdf |
| 12 | cc.WE.PD2D3.up.PWAY.pdf |

**Supplementary Data File 5** (continuation of legend)

Networks illustrating all the functional categories significantly enriched (p.adjust < 0.01) in analysis of the gene products represented by the probes significantly up- or down-regulated (adj.P.Val < 0.05) in the White European (WE) cohort treatment groups. The thickness of lines (edges) drawn to connect the nodes shown increases with increasing significance (p.adjust-values). This aimed to identify functional categories that are more extensively affected by $D_2$, or by $D_3$, or by both $D_2$ and $D_3$, than by the placebo, and took into consideration the changes occurring in the placebo treatment group during the period of the study. Network names correspond to the worksheets in the excel spreadsheet **Supplementary Data File 4** summarising the comparative functional enrichment analysis results, from which these networks were constructed.

Thus:

- Names ending GOBP contain results from Gene Ontology analysis using the Biological Process subontology.
- Names ending GOCC contain results from Gene Ontology analysis using the Cellular Compartment sub-ontology.
- Names ending GOMF contain results from Gene Ontology analysis using the Molecular Function sub-ontology.
- Names ending KEGG contain results from KEGG pathway analysis.
- Names ending PWAY contain results from Reactome pathway analysis.

- Names commencing cc.WE.PD2D3.down contain results from the analysis of lists of down-regulated genes from the White European D2, D3 or Placebo (P) treatment groups.

- Names commencing cc.WE.PD2D3.up contain results from the analysis of lists of up-regulated genes from the White European D2, D3 or Placebo (P) treatment groups.

- Names containing “down” = down-regulated in V3 compared to V1 in the treatment groups.

- Names containing “up” = up-regulated in V3 compared to V1 in the treatment groups.

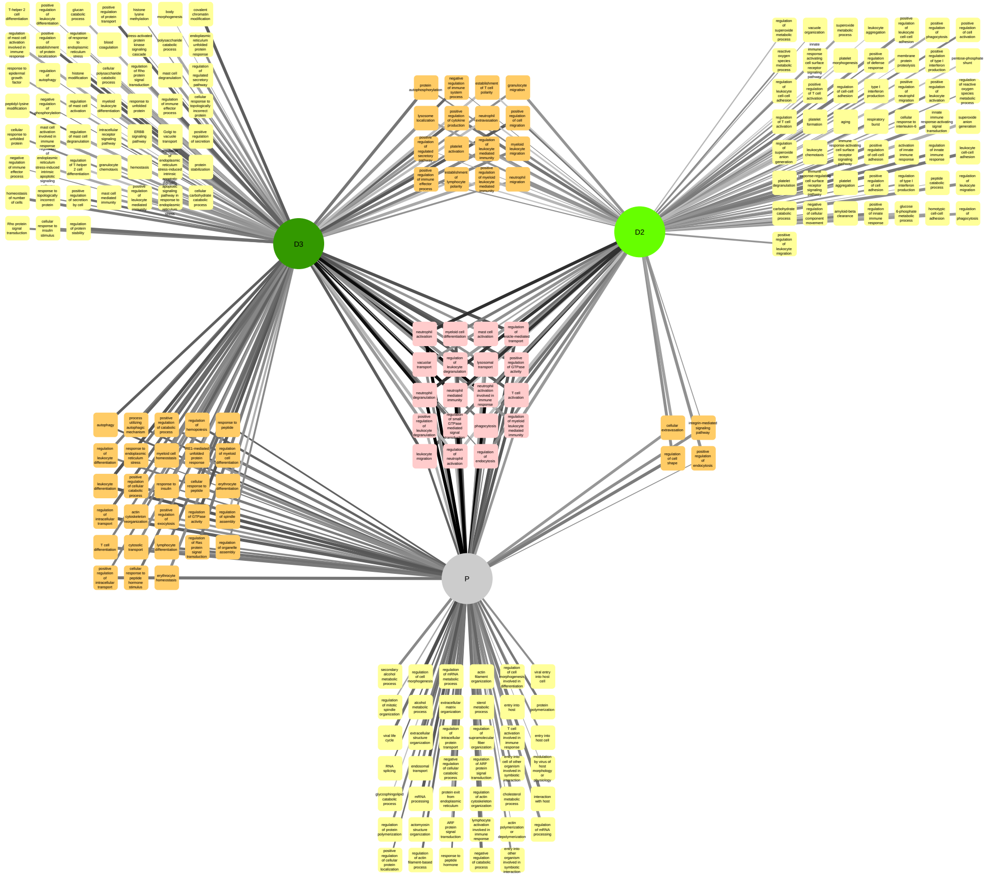


cc.WE.PD2D3.down.GOBP

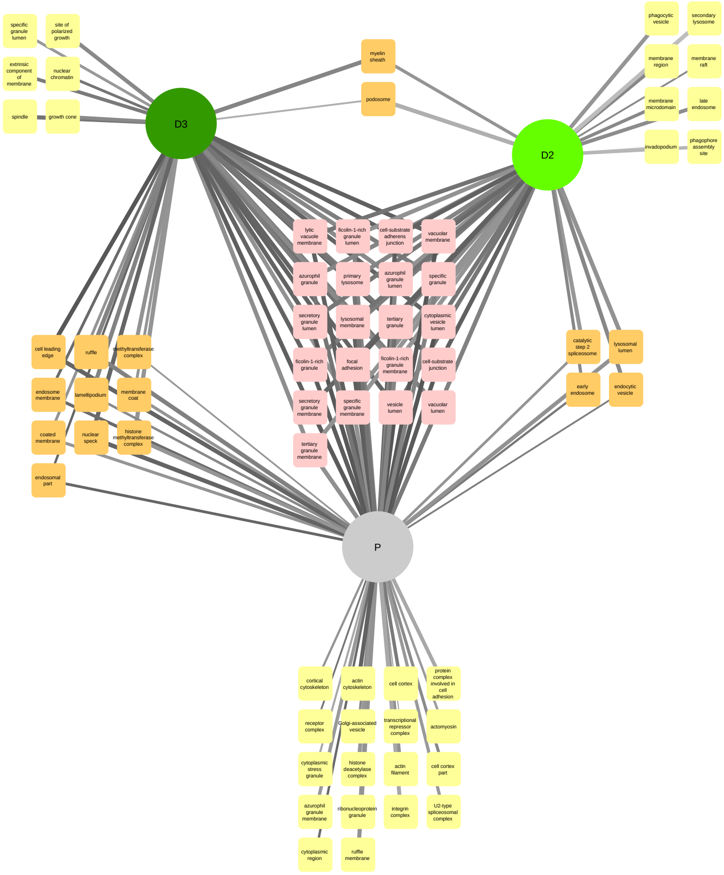


cc.WE.PD2D3.down.GOCC

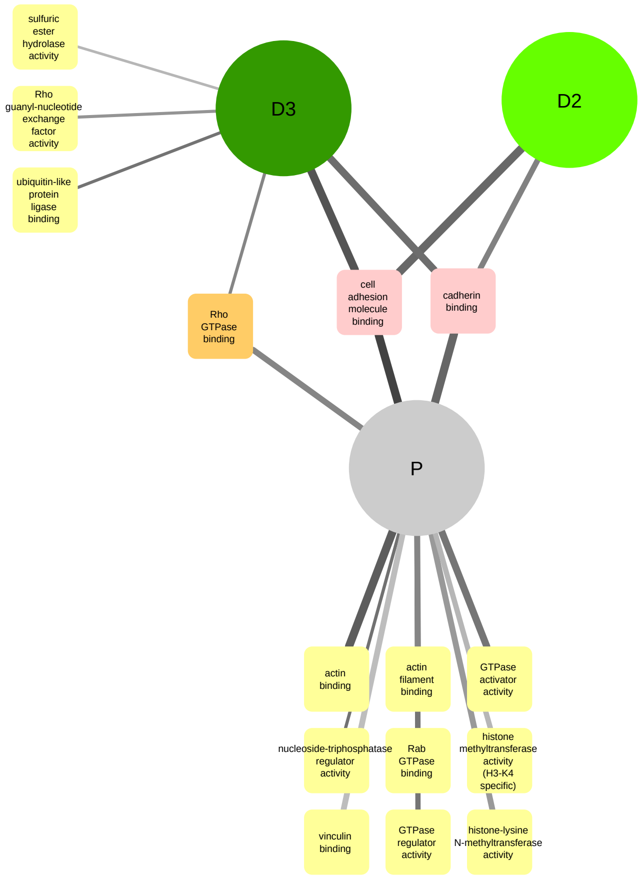


cc.WE.PD2D3.down.GOMF

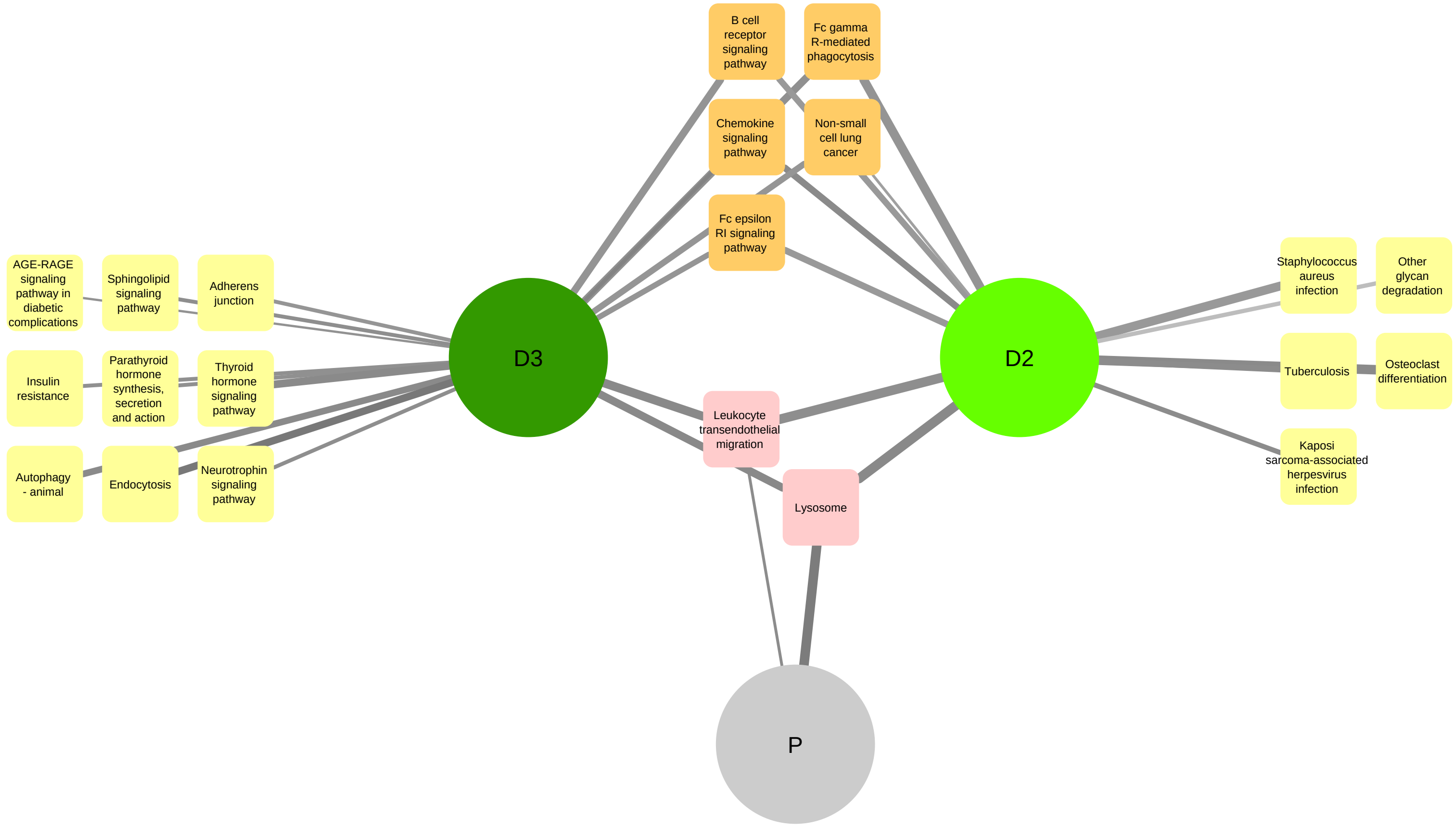


cc.WE.PD2D3.down.KEGG

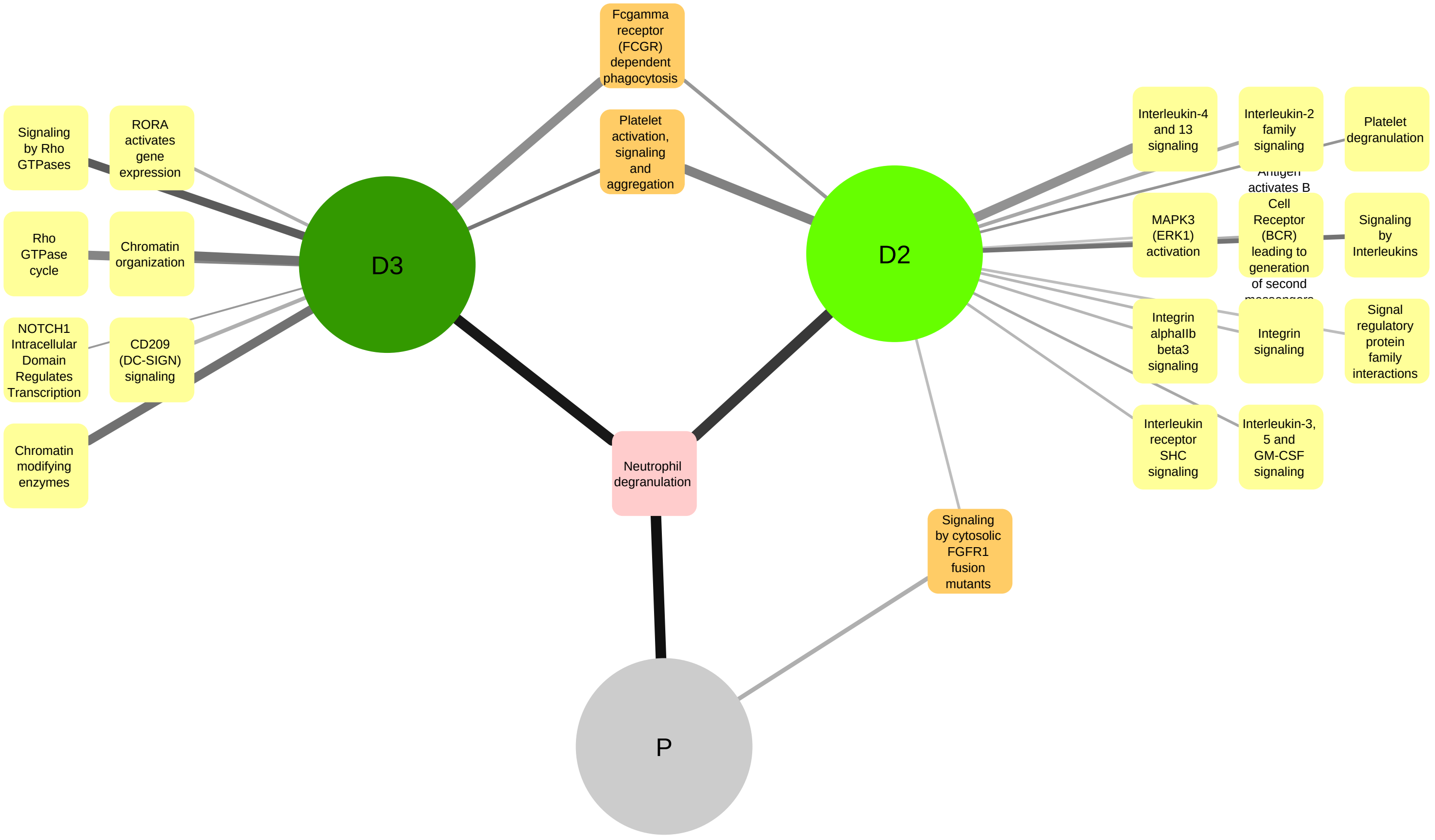


cc.WE.PD2D3.down.PWAY

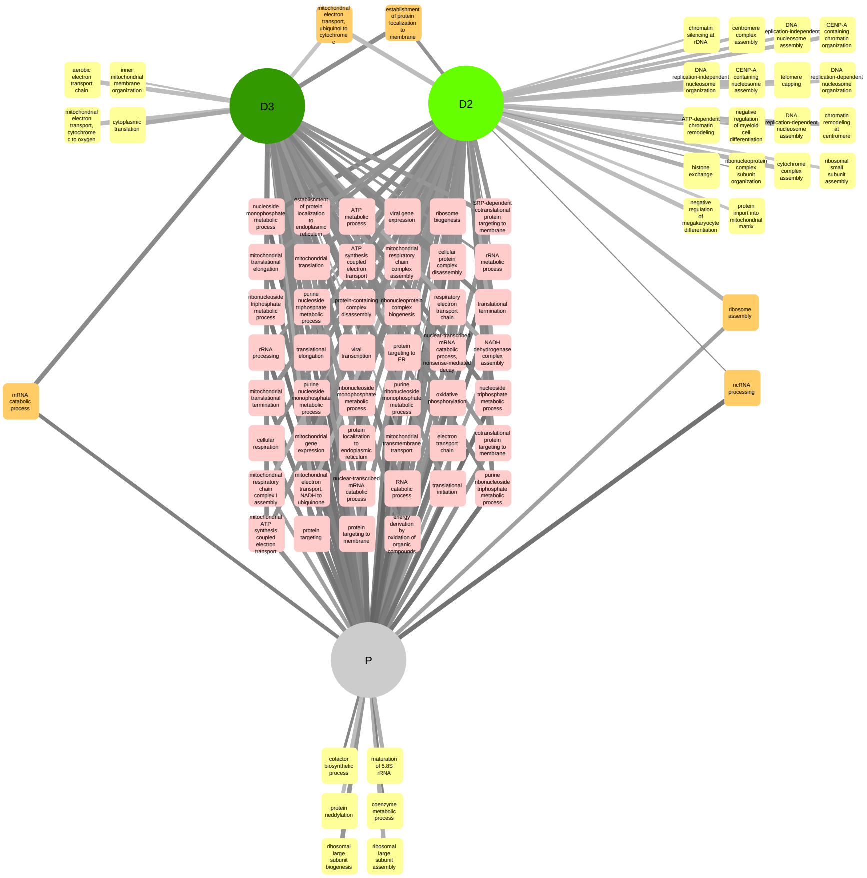


cc.WE.PD2D3.up.GOBP

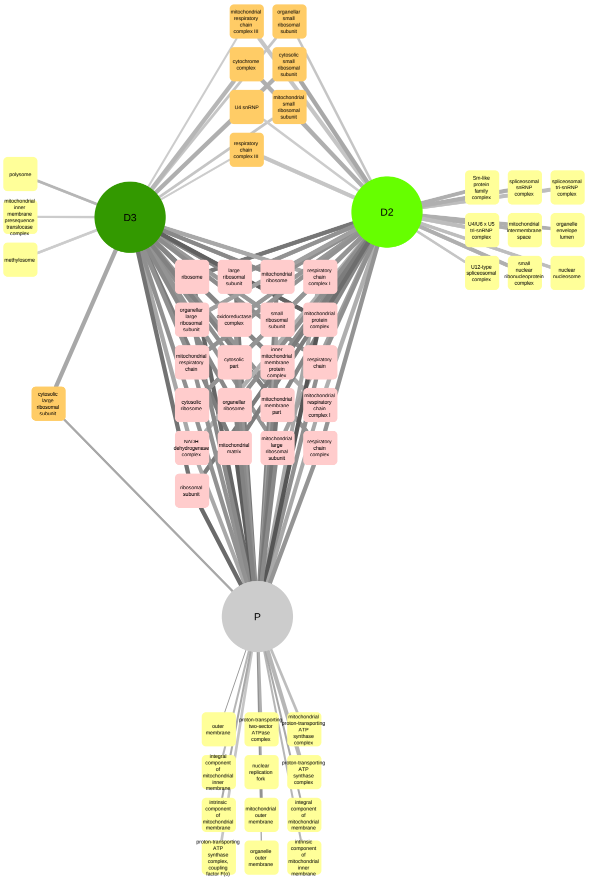


cc.WE.PD2D3.up.GOCC

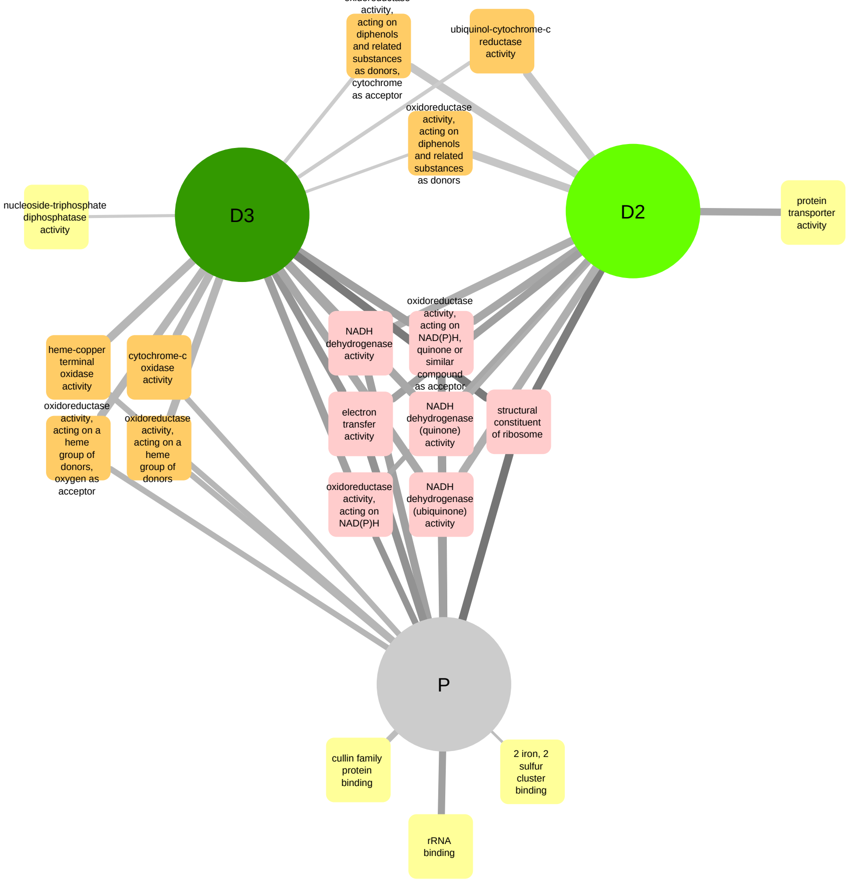


cc.WE.PD2D3.up.GOMF

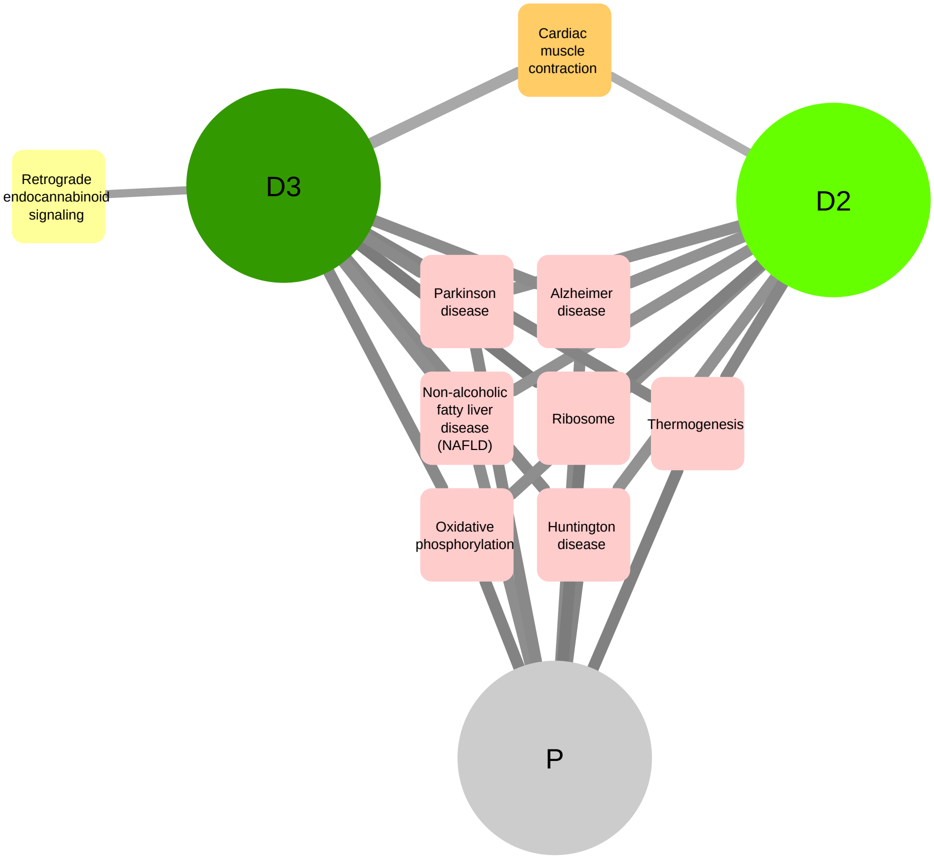


cc.WE.PD2D3.up.KEGG

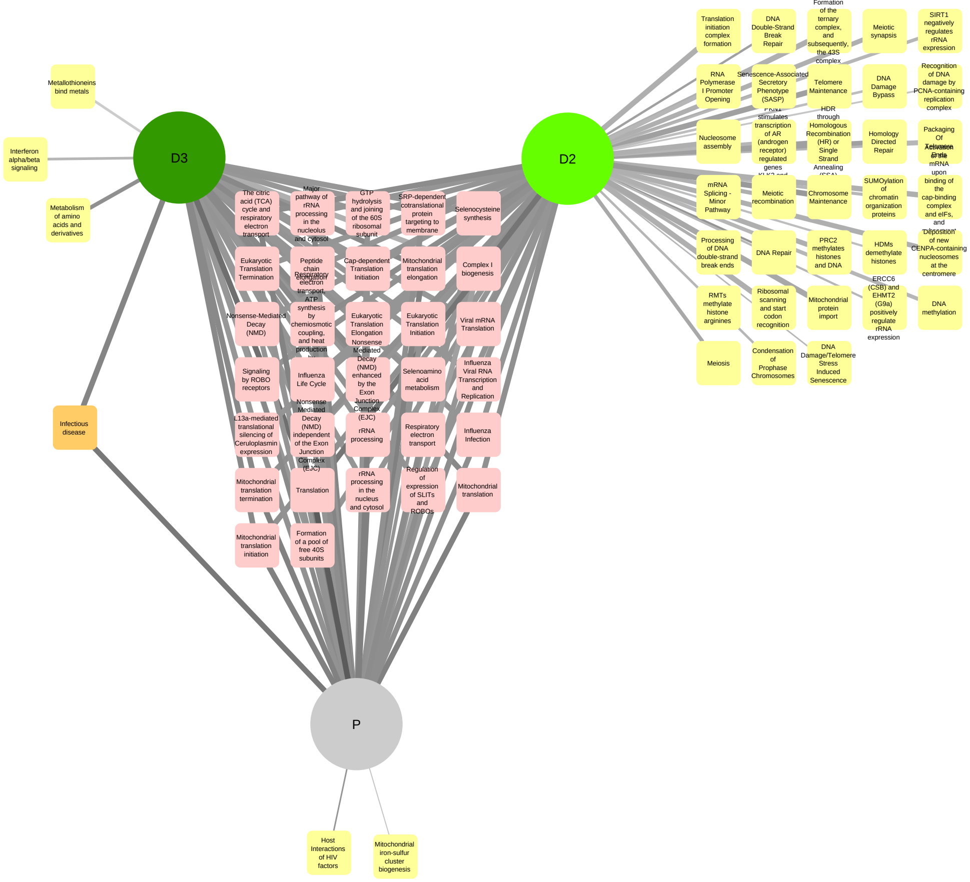


cc.WE.PD2D3.up.PWAY
